# Proxiaml Temporary Occlusion using Balloon Guide Catheter for Mechanical Thrombectomy (PROTECT-MT): Study protocol for a multicenter randomized trial

**DOI:** 10.1101/2024.07.12.24310194

**Authors:** Yu Zhou, Lei Zhang, Yongwei Zhang, Zifu Li, Pengfei Xing, Hongjian Shen, Xuan Zhu, Mayank Goyal, Jianmin Liu, Pengfei Yang, PROTECT-MT investigators

## Abstract

**Background and Rationale:** Uncertainty exists over balloon guide catheter(BGC) usage for Mechanical Thrombectomy(MT) in patients with acute ischemic stroke acute ischemic stroke due to anterior circulation large vessel occlusion.

**Objective:** To determine the effectiveness of balloon guide catheter(BGC) as compared to conventional guide catheter on functional outcome in patients with acute ischemic stroke and treated with MT.

**Design:** Proximal TEmporary oCclusion using balloon guide caTheter for Mechanical Thrombectomy (PROTECT-MT) is an investigator-initiated, multicenter, prospective, open label trial with blinded outcome assessment (PROBE design) assessing the effectiveness and safety of BGC in acute ischemic stroke. An estimated 1074 patients meet the inclusion criteria would be randomized (1:1) to receive either MT with BGC(intervention) or MT with conventional guide catheter(control). The primary outcome is an ordinal shift analysis of scores on the modified Rankin scale (mRS) at 90 days.

**Progress:** Recruitment started on February 7, 2023. At a meeting of the independent Data and Safety Monitoring Board in November 2023 to review primary outcome data available for 169 patients, they recommended suspension of recruitment due to safety concerns, which was implemented by the Trial Steering Committee (TSC) with 329 randomized patients. The TSC stopped recruitment due to the safety concerns persisted on further review of the data on April 18, 2024.

**Discussion:** PROTECT-MT is the largest RCT assessing the effectiveness of BGC versus conventional guide catheter on functional outcome in large vessel occlusion patients treated with MT, it will provide further randomized evidence on the role of BGC in AIS patients treated with MT.

## Introduction

During the procedures of Mechanical Thrombectomy(MT), devices played an important role in the success of vascular recanalization. A balloon guide catheter (BGC) is an adjunctive device used to arrest and reverse flow by inflating the balloon at its tip, which allows for flow reversal in intracranial arteries during retrieval of thrombectomy devices by applying concomitant aspiration through its lumen.^1^ Multiple in vitro and animal studies show that BGC usage reduces distal emboli in case of clot fragmentation compared to conventional guide catheters.^2^ Some observational studies have also shown that the use of BGC improved the final reperfusion quality, shorten the procedure time, and may lead to better clinical outcomes in patients undergoing thrombectomy.^3–10^ Based on these findings, BGC usage during MT is recommended or preferred in current American and European guidelines.^11,12^

However, high-level evidence data are still absent.^13^ While substantial observational pre-clinical and clinical data point towards a beneficial effect of BGCs, results of some other studies did not supported such findings.^14–18^ Some recent studies showed that BGC may be unnecessary when aspiration catheter with/without stent is used, and the rates of recanalization and functional outcome were similar between patients treated with BGC and conventional guide catheter.^14,19^ In addition, some technical issues, such as stiffness and trackability of the catheter, prolonged procedure time due to BGC preparing and delivering, and restricted compatibility with larger bore aspiration catheters et al, as well as increased cost, hindered the widespread use of BGCs.^14^ These issues aroused debates on whether BGC should be routinely used in thrombectomy treatment.^13^ In an international survey, only 1 out of 4 neurointerventionalists routinely use a BGC when performing MT.^20^

Due to these practice heterogeneities and lack of high-level evidence data, we initiated PROTECT-MT trial to assess the effectiveness and safety of BGC usage in MT for acute ischemic stroke due to large vessel occlusion.

## Methods

### Study Aims

To determine the effectiveness of balloon guide catheter(BGC) as compared to conventional guide catheter on functional outcome (modified Rankin Scale [mRS] score) in patients with acute ischemic stroke due to anterior circulation large vessel occlusion.

### Study design

This is a prospective, multicenter, randomized controlled, open-label, blinded outcome assessment trial involving eligible subjects to be recruited from 40-60 hospitals in China. Subjects who meet the eligibility criteria will be randomized to either intervention group (receiving treatment with BGC) or control group (receiving treatment with conventional guide catheter) (**Figure 1**). Followed up would be conducted at 1 day, 7 days, and 90 days after thrombectomy (**Table 1**).

**Figure 1.**
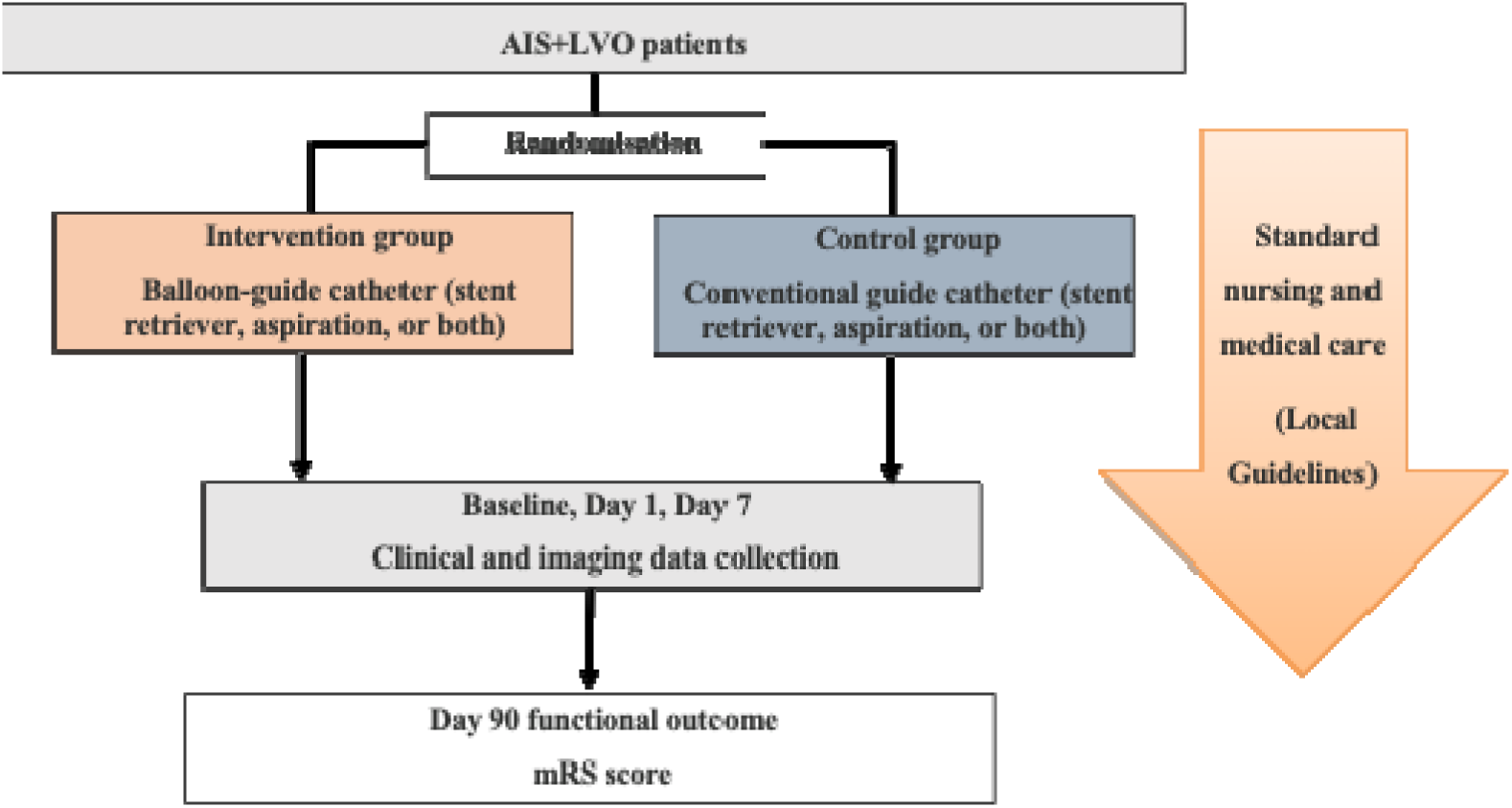
Flow chat in PROTECT-MT. AIS denotes acute ischemic stroke. LVO denotes large vessel occlusion. mRS denotes modified Rankin Scale.

**Table 1.**
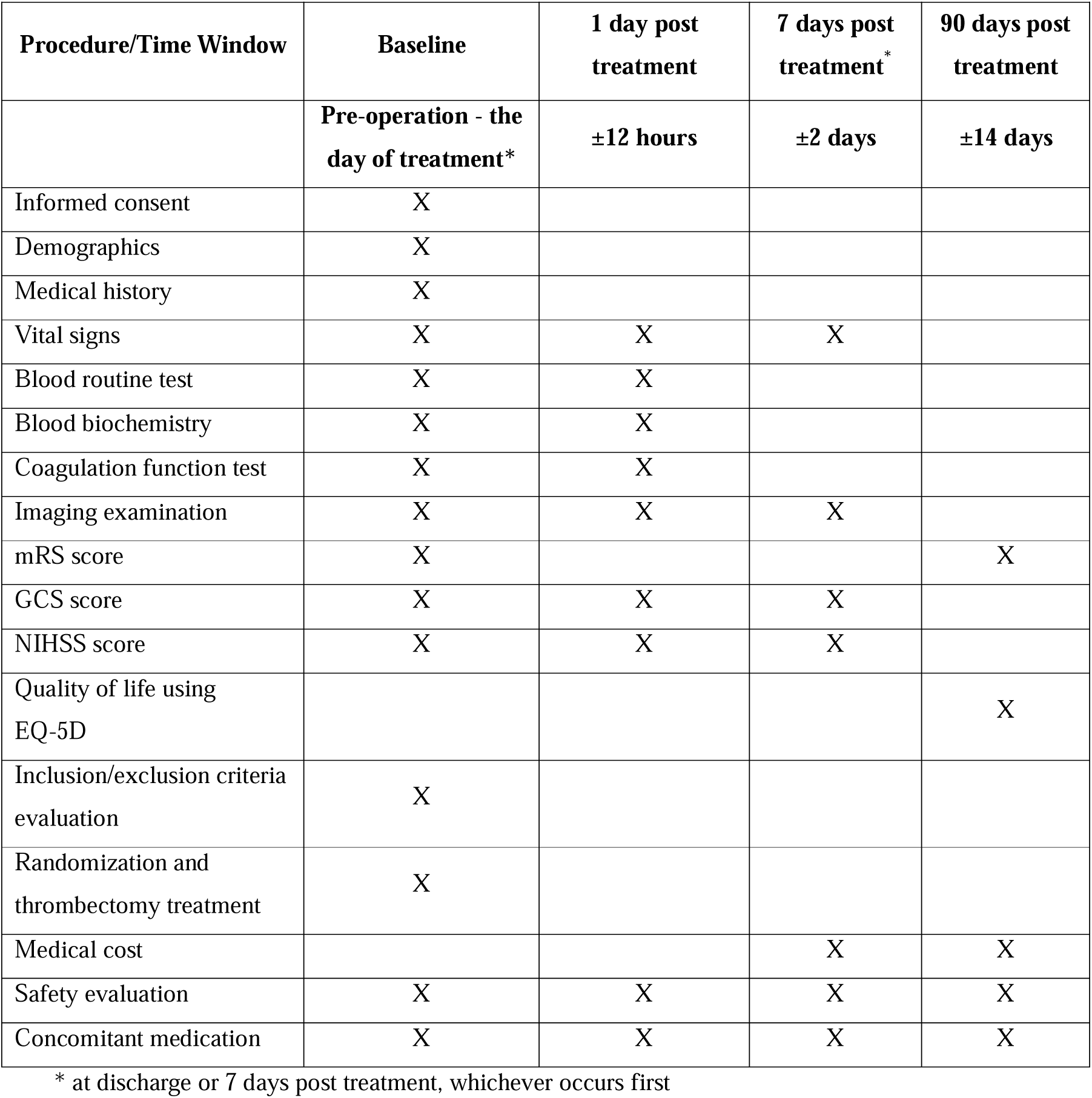
Timing of all procedures in PROTECT-MT.

Consent is obtained upon a patient’s presentation to hospital prior to proceeding to cerebral angiography, with randomization undertaken after confirming the inclusion and exclusion criteria. The study is conducted in compliance with local and international regulatory and ethical requirements. The medical ethical committee and research board of the Changhai Hospital, Shanghai, approved this study in China (CHEC2023-301).

### Population

Subjects with acute ischemic stroke(AIS) due to due to anterior circulation large vessel occlusion(LVO) confirmed by cranial imaging and are eligible for MT according to local guidelines will be enrolled into this study. The principal investigator at each site will be responsible for subject screening and enrollment. Detailed inclusion and exclusion criteria are shown as below.

Inclusion Criteria: 1)Age ≥ 18 years; 2)Diagnosis of AIS with confirmed anterior circulation LVO (including intracranial segment of the internal carotid artery, and the first or proximal second segment [M1 or proximal M2] of the middle cerebral artery) by brain imaging; 3)To receive MT within 24 hours after AIS onset according to local guidelines; 4)Preoperative mRS score of 0-1; 5)Signed informed consent form obtained from the subject (or approved surrogate).

Exclusion Criteria: 1) Intracranial hemorrhage confirmed by imaging; 2)Known or suspected pre-existing (chronic) large vessel occlusion in the symptomatic territory; 3)Excessive vascular access tortuosity disables the use of balloon guide catheter; 4)Intracranial stent implanted in the symptomatic territory that precludes the deployment/removal of the mechanical thrombectomy device; 5) Any other condition that precludes the performing of mechanical thrombectomy procedure; 6)Occlusions in multiple vascular territories confirmed by Computed Tomography Angiography(CTA) or Magnetic Resonance Angiography(MRA); 7)Subjects who are pregnant;8)Subjects who are allergy to the contrast agent; 9) Subjects who refuse to cooperate or unable to tolerate interventional operation;10) Subjects whose expected lifetime are less than 90 days; 11) Subjects who are unlikely to participate in follow-up assessments according to the investigator’s judgement; 12) Any other condition that, according to the investigator’s judgement, not suitable for using of balloon guide catheter.

### Randomisation and blinding

Subjects will be randomized via an Internet-based randomization system in a 1:1 manner to treatment with BGCs (Intervention group) or not(control group). The randomization sequence will use a minimization algorithm to ensure balance in key prognostic factors, according to stratifying variables of site of recruitment, preferred thrombectomy strategy (stent retriever VS aspiration VS stent retriever + aspiration) and time from symptom onset to recanalization (< 6 vs 6 h).

Both patient and treating physician will be aware of the treatment assignment. But outcome evaluation would be blinded. Information on outcome at 90 days will be assessed through standardized forms and procedures, by a trained investigator blinded for treatment allocation. Interviews will be recorded. Assessors who are blinded to the treatment allocation will perform assessment of outcome on the modified Rankin scale on this information. Neuro-imaging will be also assessed by a core-laboratory blinded for treatment allocation. Information on treatment allocation will be kept separate from the main study database. The steering committee will be kept unaware of the results of interim analyses of efficacy and safety. An independent DSMB statistician will combine data on treatment allocation with the clinical data to report to the data monitoring committee (DSMB).

### Intervention

Subjects will be treated with thrombectomy based on the results of randomization. Subjects in intervention group would be treated with BGCs combined with conventional thrombectomy, while subjects in control group will be treated with conventional guide catheter combined with conventional thrombectomy.

Any National Medical Products Administration (NMPA) devices (including the use of a thrombectomy stent, aspiration catheter, or combined use of both) are allowed.

### Outcome

#### Primary outcome

The primary outcome is patient functional outcome, defined as modified Rankin Scale (mRS) score shift, at 90 days.

#### Secondary Outcomes

1)Dichotomized mRS at 90 days after the procedure (0-1 versus 2-6, 0-2 versus 3-6,0-3 versus 4-6, 0-4 versus 5-6, 0-5 versus 6); 2)Change in stroke severity (NIHSS score) at 24 hours post treatment;3)Change in stroke severity (NIHSS score) at 7 days post treatment or discharge (whichever occurs first);4)Final infarction volume at 5-7 days post treatment; 5)Technical success rate (defined as successfully navigating the guide catheter into the target vessel, and finishing the mechanical thrombectomy procedure without changing to another guide catheter); 6)Reperfusion outcome (eTICI 2b or greater, eTICI 2c or greater, eTICI 3) in final angiogram; 7)Reperfusion outcome (eTICI 2b or greater, eTICI 2c or greater, eTICI 3) after the first pass; 8)Time from groin puncture to successful reperfusion (eTICI 2b or greater, eTICI 2c or greater); 9)Percentage of subjects with acceptable revascularization quality (eTICI 2b or greater, eTICI 2c or greater) within 45 min of access; 10)Number of thrombectomy attempts (final); 11)Occurrence of emboli to a new territory.

#### Safety outcomes

1)Deaths at 90 days (±14 days) post treatment; 2) Intracranial hemorrhage, symptomatic intracranial hemorrhage or asymptomatic intracranial hemorrhage at 7 days post treatment or discharge (whichever occurs first); 3) Other serious adverse events (SAEs) adjudicated by the Clinical Events Committee; 4) Any peri-procedural complications, including vessel dissection, arterial perforation, femoral access complications, contrast allergy, etc.

### Cost Outcomes

1)Health-related quality of life using EQ-5D; 2)Utility-weighted mRS scores; 3)Duration of hospitalization; 4)Treatment cost.

### Data and Safety Monitoring Board

An independent expert Data and Safety Monitoring Board (DSMB) is responsible for reviewing the safety, ethics, and accumulating data for the study. The members are governed by a charter outlining their responsibilities, procedures, and confidentiality, for monitoring the efficacy and safety out-comes for early dramatic benefits or potential harmful effects, and to provide reports to the Trial Steering Committee (TSC) with recommendations to continue, pause, or terminate recruitment. They review unblinded data on recruitment, adherence to the protocol, primary and secondary outcomes, and serious adverse events (SAEs), at regular intervals.

### Sample Size

In a meta-analysis comparing BGC with conventional guide catheter, the use of BGC improved the proportion of functional outcome by 10.8%.^21^ In this study, based on a more conservative estimate, we assume the usage of BGC would improve the function outcome by 8%.

Based on the distribution of the mRS in the control group of the trial, which we derived from the intervention group of the MR CLEAN trial^22^: mRS0: 3%; mRS 1: 9%; mRS 2: 21%; mRS 3: 18%; mRS 4: 22%; mRS 5: 6% and mRS 6:21%. We assumed a favorable treatment effect with a common odds ratio (cOR) of 1.43, corresponding to an 8% absolute increase in the rate of mRS scores of 0-2. In a simulation with 5000 runs we computed the proportion of positive trials, for a given sample size. A sample size of 1074 subjects is estimated to be able to demonstrate this treatment effect with 87% power and 5% type-1 error. This sample size also allows for 5% dropout rate and 5% crossover rate.

### Statistical analysis

Analyses will be performed based on the intention-to-treat (ITT) principle. Baseline data by treatment allocation will be reported with statistical procedures. Missing baseline characteristics will be imputed using regression imputation as appropriate. The primary endpoint will be analyzed by means of an ordinal logistic regression. Pre-defined subgroups will be analyzed by testing for interaction between the specific baseline characteristic and treatment. Any reported SAE or discontinuation of assigned treatment due to an SAE will be classified using MedDRA. Statistical analyses will be detailed in the pre-specified Statistical Analysis Plan (SAP).

The study consists of 2 interim analyses to be performed when 30% and 60% of the 90-day follow-up data have been collected. The independent data and safety monitoring board (DSMB) will adopt the Haybittle-Peto rule, and α <0.001 in the interim analysis will be considered as statistically significant. Because 2 interim analyses will be performed, the significance level of α = 0.0482 will be used for final analysis. The DSMB will monitor SAEs (e.g. death, spontaneous intracerebral hemorrhage, and neurological deterioration) periodically, and the occurrence of excessive SAEs will trigger discussion regarding termination of the study.

### Progress to date

Recruitment started on February 7, 2023, in China. At a meeting of the DSMB on November 13, 2023, there were 329 patients randomized at 28 active sites in China. After reviewing primary outcome data available for 169 participants, they recommended suspension of recruitment due to safety concerns. Due to persistent concerns raised by the DSMB on further review of all available outcome data on April 18, 2024, the TSC stopped patient recruitment.

## Discussion

PROTECT-MT is a randomized clinical trial with a PROBE design assessing the effectiveness of BGC versus conventional guide catheter in LVO-AIS patients treated with MT.

Compared with other trials evaluating the role of BGC in MT,^23^ our trial has some differences in the trial design, including inclusion/exclusion criteria, control arm setting, primary outcomes, et al. To better understand the effectiveness of BGC in the real world and allow for generalization of the study results, we set conventional guide catheter, rather BGC without inflation, as control arm, and we included a wide range of population. Essentially all patients eligible for MT with an anterior circulation occlusion according to local guidelines can be included in the trial. In addition, we set the 90-day mRS as primary outcome rather other surrogate marker to direct evaluating the impact of BGC on patient outcome, which resulted in a large sample size of our trial. The meta-analysis comparing BGC with conventional guide catheter suggested, the use of BGC improved the proportion of functional outcome by 10.8%.^21^ In this study, based on a more conservative estimate, we assume the usage of BGC would improve the function outcome by 8%. Based on that, a sample size of 1074 subjects was calculated to demonstrate this treatment effect with 87% power and 5% type-1 error, which also allows for 5% dropout rate and 5% crossover rate.

Current reports indicated the effect of BGC may differ with primary techniques, while BGC seems effective in patients treated with stent retriever, controversies existed in patients treated with other techniques such as primary aspiration, and primary combined approaches (stent-retriever with distal aspiration).^14,15^ In order to minimize the risk of bias related to differences in thrombectomy techniques in the control and intervention arms, we set the primary thrombectomy technique to ensure equal and balanced distribution of the three techniques in the BGC arm and the control arm.

## Summary and conclusions

PROTECT-MT is the largest RCT assessing the effectiveness of BGC as compared to conventional guide catheter on functional outcome in patients with acute ischemic stroke due to anterior circulation large vessel occlusion. This trial will provide more reliable evidence and guide the clinical practice. Patient recruitment has now stopped, and we are preparing statistical analysis plan for further analysis.

## Supporting information

SAP

protocol version 4.0

## Data Availability

All data produced in the present study are available upon reasonable request to the authors

## Clinical trial registration

ClinicalTrials.gov Identifier: NCT05592054

