## Supplementary material for "Proxiaml Temporary Occlusion using Balloon Guide Catheter for Mechanical Thrombectomy (PROTECT-MT): Study protocol for a multicenter randomized trial": SAP

**Statistical Analysis Plan:**

| | |
|---|---|
| Protocol Number: | PROTECT-MT |
| SAP Version / Status (Draft / Final / Amendment): | 1.0 / Final |
| Date: | Jul 8th, 2024 |

### PROximal TEmporary oCclusion using balloon guide caTheter for Mechanical Thrombectomy (PROTECT-MT)

**Trial Statistician (CRO):**

Sam Zhong

Shanghai KNOWLANDS MedPharm Consulting Co., Ltd.

**Sponsor:**

Changhai Hospital Affiliated to Naval Medical University

### Signature Page

**PROximal TEmporary oCclusion using balloon guide caTheter for Mechanical Thrombectomy (PROTECT-MT)**

**Trial Statistician (CRO)**

Sam Zhong — July 8, 2024

Sam Zhong
Shanghai KNOWLANDS
MedPharm Consulting Co., Ltd.

Date

### Signature Page

#### PROximal TEmporary oCclusion using balloon guide caTheter for Mechanical Thrombectomy (PROTECT-MT)

**Sponsor**

[signature] 2024.7.8

Pengfei Yang
Changhai Hospital Affiliated to
Naval Medical University

Date

### LIST OF ABBREVIATIONS AND DEFINITIONS OF TERMS

| Abbreviation | Definition of terms |
|---|---|
| AE | Adverse Event |
| AIS | Acute Ischemic Stroke |
| BGC | Balloon Guide Catheter |
| CI | Confidence Interval |
| CRF | Case Report Form |
| DSA | Digital Subtraction Angiography |
| DSMB | Data and Safety Monitoring Board |
| GCS | Glasgow Coma Scale Score |
| EQ-5D | EuroQol-5 Dimensions |
| eCRF | Electronic Case Report Form |
| eTICI | Extended Thrombolysis in Cerebral Infarction |
| ITT | Intention-to-Treat Population |
| LVO | Large Vessel Occlusion |
| MedDRA | Medical Dictionary for Drug Regulatory Activities |
| MRI | Magnetic Resonance Imaging |
| mRS | Modified Rankin Scale |
| NIHSS | National Institute of Health stroke scale |
| OR | Odds Ratio |
| PPS | Per-protocol Set |
| PT | Preferred Term |
| SAE | Serious Adverse Event |
| SAP | Statistical Analysis Plan |
| SOC | System Organ Class |

### 1 INTRODUCTION

This Statistical Analysis Plan (SAP) is developed based on the most recent study protocol (Version 4.0, 25-Oct-2023) and electronic Case Report Form (eCRF, Version 1.2, 07-Nov-2023), and details the statistical analysis strategies and methods for the study.

The SAP predefines the statistical analysis population, analysis variables and analysis methods before database lock to ensure the reliability of the study results.

#### 1.1 Study Design

This is a prospective, multicenter, randomized controlled, open-label, blinded outcome assessment trial involving 1,074 eligible subjects with acute ischemic stroke (AIS) due to anterior circulation large vessel occlusion (LVO) to be recruited from 40-60 hospitals in China. Subjects who meet the eligibility criteria will be randomized to either intervention group (receiving treatment with balloon guide catheter [BGC]) or control group (receiving treatment with conventional guide catheter). Follow up visits will be conducted at 1 day, 7 days, and 90 days after thrombectomy. Study design is presented as below (Figure 1):

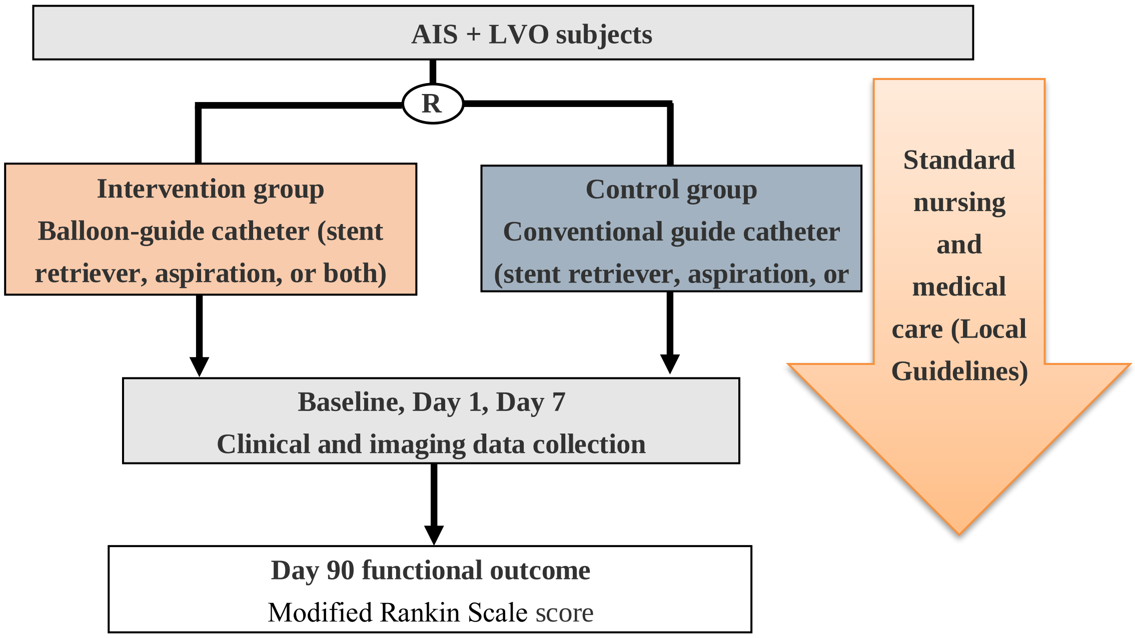

**Figure 1. Study schema**

#### 1.2 Study Objectives

The objective of this study is to determine the effectiveness of BGC as compared to conventional guide catheter on functional outcome (modified Rankin Scale [mRS] score) in patients with acute ischemic stroke due to anterior circulation large vessel occlusion.

#### 1.3 Study outcomes

**Primary efficacy outcome:**

- The primary outcome is functional outcome, defined as the mRS score shift at 90 days (± 14 days).

**Secondary efficacy outcomes:**

- Dichotomized mRS at 90 days after the procedure (0-1 vs. 2-6, 0-2 vs. 3-6, 0-3 vs. 4-6, 0-4 vs. 5-6, 0-5 vs. 6);
- Change in stroke severity (The National Institutes of Health Stroke Scale [NIHSS] score, the criteria for scoring are detailed in Appendix 1) at 24 hours post treatment;
- Change in stroke severity (NIHSS score) at 7 days post treatment or discharge (whichever occurs first);
- Final infarction volume;
- Technical success rate (defined as successfully navigating the guide catheter into the target vessel, and finishing the mechanical thrombectomy procedure without changing to another guide catheter);
- Reperfusion outcome (Extended Treatment In Cerebral Ischemia [eTICI] 2b or greater, eTICI 2c or greater, eTICI 3; eTICI criteria are detailed in Appendix 2) in final angiogram;
- Reperfusion outcome (eTICI 2b or greater, eTICI 2c or greater, eTICI 3) after the first pass;
- Time from groin puncture to successful reperfusion (eTICI 2b or greater, eTICI 2c or greater);
- Percentage of subjects with acceptable revascularization quality (eTICI 2b or greater, eTICI 2c or greater) within 45 min of access;
- Number of thrombectomy attempts (final) ;
- Occurrence of emboli to a new territory.

**Safety outcomes:**

- Deaths at 90 days (±14 days) post treatment;
- Intracranial hemorrhage, symptomatic intracranial hemorrhage or asymptomatic intracranial hemorrhage at 7 days post treatment or discharge, whichever occurs first;
- Other serious adverse events (SAEs) adjudicated by the Clinical Events Committee;
- Any peri-procedural complications, including vessel dissection, arterial perforation, and femoral access complications, etc.

**Cost Outcomes:**

- Health-related quality of life using EuroQol-5 Dimension [EQ-5D] (Appendix 3);
- Utility-weighted mRS scores;

- Duration of hospitalization;
- Treatment cost.

#### 1.4 Estimation of Sample Size

In a meta-analysis [1] comparing BGC with conventional guide catheter, the use of balloon guide catheter improved the proportion of functional outcome by 10.8%. In this study, based on a more conservative estimate, we assume the usage of BGC would improve the function outcome by 8%. Based on the distribution of the mRS in the control group of the trial, which we derived from the intervention group of the MR CLEAN trial : mRS0: 3%; mRS 1: 9%; mRS 2: 21%; mRS 3: 18%; mRS 4: 22%; mRS 5: 6% and mRS 6:21%. We assumed a favorable treatment effect with a common odds ratio (cOR) of 1.43, corresponding to an 8% absolute increase in the rate of mRS scores of 0-2. In a simulation with 5000 runs we computed the proportion of positive trials, for a given sample size. A sample size of 1074 subjects is estimated to be able to demonstrate this treatment effect with 87% power and 5% type-1 error. This sample size also allows for 5% dropout rate and 5% crossover rate.

#### 1.5 Randomization and Treatment allocation

Subjects will be randomized via an internet-based randomization system in a 1:1 manner to treatment with BGCs (Intervention group) or not (Control group), stratified by site, preferred initial thrombectomy strategy (stent retriever vs. aspiration vs. stent retriever + aspiration), and onset to randomization time (<6 hours vs. ⩾6 hours). The randomization sequence will use a minimization algorithm to ensure balance in these key prognostic factors. Randomization is allowed when large vessel occlusion has been established.

Subjects will be treated with thrombectomy based on the results of randomization:

- **Intervention group**: Subjects treated with BGCs combined with conventional thrombectomy.
- **Control group**: Subjects treated with conventional guide catheter combined with conventional thrombectomy.

Any National Medical Products Administration (NMPA) approved devices (including thrombectomy stent, aspiration catheter, or combined use of both) are allowed in this study.

#### 1.6 Study Procedure

Before starting the study, patients or their guardians must read and sign the informed consent approved by the current Ethics Committee (EC). All research steps should be carried out within the time window specified in the study protocol.

The study consists of a total of 4 visits per subject, including: visits at baseline (pre-procedure ~ the day of treatment), 1 day (±12 hours) post treatment, 7 days (±2 days) post treatment or at discharge, and 90 days (±14 days) post treatment.

The study procedure is shown in the table 1 below.

**Table 1. Schedule of the study procedures**

| Procedure/Time Window | Baseline | 1 day post treatment | 7 days post treatment# | 90 days post treatment |
|---|---|---|---|---|
| | Pre-operation - the day of treatment* | ± 12 hours | ± 2 days | ± 14 days |
| Informed consent | X | | | |
| Demographics | X | | | |
| Medical history | X | | | |
| Vital signs | X | X | X | |
| Blood routine test | X | X | | |
| Blood biochemistry | X | X | | |
| Coagulation function test | X | X | | |
| Imaging examination | X | X | X | |
| mRS score | X | | | X |
| GCS score | X | X | X | |
| NIHSS score | X | X | X | |
| Quality of life using EQ-5D | | | | X |
| Inclusion/exclusion criteria evaluation | X | | | |
| Randomization and thrombectomy treatment | X | | | |
| Medical cost | | | X | X |
| Safety evaluation | X | X | X | X |
| Concomitant medication | X | X | X | X |

Notes: #: at discharge or 7 days post treatment, whichever occurs first. *: The end date of surgery will be considered as the day of surgery.

### 2 STATISTICAL ANALYSIS METHODOLOGY

#### 2.1 Statistical Analysis Variables

This statistical analysis plan (SAP) will include variables to be analyzed as follows: demographics and baseline characteristics, treatment efficacy and safety data.

##### 2.1.1 Demographics and baseline characteristics

The demographic and baseline information will include age (years), sex, nationality, height (cm), weight (kg), medical history, smoking and drinking history, baseline medications, imaging examination, mRS score, NIHSS score and GCS score.

##### 2.1.2 Efficacy outcomes

###### 2.1.2.1 Primary efficacy outcome– mRS score

The modified Rankin Scale (mRS) is an ordinal hierarchical scale ranging from 0 to 5, with higher scores indicating more severe disability. In addition, a score of 6 is added to signify death; the criteria for scoring are detailed in Table 2:

**Table 2. Modified Rankin Scale**

| Score | Definition |
|---|---|
| 0 | No symptoms |
| 1 | Symptoms, no disability: Minor symptoms that do not interfere with lifestyle |
| 2 | Slight disability: Slight disability, symptoms that lead to some restriction in lifestyle, but do not interfere with the patient's capacity to look after himself |
| 3 | Moderate disability: Moderate disability, symptoms that significantly restrict lifestyle and prevent totally independent existence |
| 4 | Moderately severe disability: Moderately severe disability, symptoms that clearly prevent independent existence though not needing constant attention |
| 5 | Severe disability, totally dependent patient requiring constant attention day and night |
| 6 | Death |

The mRS score collected at 90 ± 14 days post treatment will be blindly evaluated by an independent Outcome Assessment Committee.

###### 2.1.2.2 Secondary efficacy outcomes

- Dichotomized mRS of 0-1 vs. 2-6 at 90 days (± 14 days);
- Dichotomized mRS of 0-2 vs. 3-6 at 90 days (± 14 days);
- Dichotomized mRS of 0-3 vs. 4-6 at 90 days (± 14 days);

- Dichotomized mRS of 0-4 vs. 5-6 at 90 days (± 14 days);
- Dichotomized mRS of 0-5 vs. 6 at 90 days (± 14 days);
- Change in stroke severity (NIHSS score) at 24 hours post treatment;
- Change in stroke severity (NIHSS score) at 7 days post treatment or discharge, whichever occurs first;
- Final infarction volume (mL);
- Technical success rate;
- Reperfusion outcome(eTICI 2b or greater, eTICI 2c or greater, eTICI 3) in final angiogram;
- Reperfusion outcome(eTICI 2b or greater, eTICI 2c or greater, eTICI 3) after the first pass;
- Time from groin puncture to successful reperfusion(eTICI 2b or greater, eTICI 2c or greater);
- Percentage of subjects with acceptable revascularization quality (eTICI 2b or greater, eTICI 2c or greater) within 45 min of access;
- Number of thrombectomy attempts (final)
- Occurrence of emboli to a new territory

**NIHSS Scale**

The NIHSS is an ordinal hierarchical scale used to evaluate the severity of stroke by assessing the patient's physical strength. Scores may range from 0 to 42, with higher scores indicating more severe ischemia. Scale items includes level of consciousness, level of consciousness questions, loc commands, best gaze, visual, facial palsy, motor arm, motor leg, limb ataxia, sensory, best language, dysarthria, extinction and inattention (the criteria for scoring are detailed in Appendix 1).

NIHSS scores assessment will be collected at baseline, 24±12 hours and 7±2 days post treatment.

**eTICI Scale**

The eTICI classification includes 0, 1, 2a, 2b, 2c and 3 (eTICI criteria are detailed in Appendix 2).

eTICI grading will be collected before and after treatment.

**Technical success rate**

The technical success rate is defined as successfully navigating the guide catheter into the target vessel, and finishing the mechanical thrombectomy procedure without changing to another guide catheter.

##### 2.1.3 Safety

###### 2.1.3.1 Adverse events (AEs)

This study focused on the SAEs and peri-procedural complications adjudicated by the Clinical Events Committee.

All SAEs terms will be coded using MedDRA 25.0 or higher before study database lock. MedDRA System Organ Class (SOC) and Preferred Term (PT) will be presented in the summary tables.

**Definition of SAEs**

Any SAEs that occurs during the study will be recorded by the investigator from the time that the study treatment is started for the subject.According to the WHO International Centre for Drug Monitoring (1994), SAEs is defined as any of the following untoward medical events which:

1) Results in death;
2) Is life threatening in the view of the principal investigator (i.e. its occurrence places the patient or subject at immediate risk of death. It does not include an adverse event, had it occurred in a more severe form, might have caused death);
3) Requires inpatient hospitalization or prolongation of existing hospitalization;
4) Results in a persistent or significant disability or incapacity;
5) Result in congenital anomaly or birth defects;
6) Important medical events that may not result in death, be life-threatening, or require hospitalizations but may jeopardize the subject and may require medical or surgical intervention to prevent one of the outcomes listed above according to the principal investigator.

**Classification by relationship of SAEs to the study device/procedure**

Relationship between SAEs and study device/procedure, the investigator will evaluate the possible association between adverse events and study device/procedure according to the time relationship and its clinical judgment in terms of "definitely related", "possibly related", "definitely unrelated", "unlikely related" and "unable to determine". Among which, events whose causal relationship is judged as "definitely related" or "possibly related" will be considered as related to the study device or procedure. If a relationship to the study device or procedure was missing, the analysis was considered to be related to the study device or procedure when appropriate.

**Definition of peri-procedural complications**

Peri-procedural complications for this study refers to complications at time of discharge or 7 days post-randomization, which include procedural complications, procedure related complications, etc., such as vessel dissection, arterial perforation, and femoral access complications.

**Classification by relationship of peri-procedural complications to the study device/procedure**

Relationship between peri-procedural complications and study device/procedure, the investigator will evaluate the possible association between adverse events and study device/procedure according to the time relationship and its clinical judgment in terms of "definitely related", "possibly related", "definitely unrelated", "unlikely related" and "unable to determine". Among which, events whose causal relationship is judged as "definitely related" or "possibly related" will be considered as related to the study device or procedure. If a relationship to the study device or procedure was missing, the analysis was considered to be related to the study device or procedure when appropriate.

###### 2.1.3.2 Vital signs

The following vital signs measurement will be collected and recorded in the eCRF: systolic blood pressure (mmHg), diastolic blood pressure (mmHg), heart rate (beat/min) and body temperature (℃).

They will be collected at baseline, 24 ± 12 hours post treatment and 7 ± 2 days post treatment.

###### 2.1.3.3 Laboratory test

**Blood routine:** red blood cells count ($*10^{12}$/L), white blood cells count ($*10^{9}$/L), neutrophils count ($*10^{9}$/L), lymphocytes count ($*10^{9}$/L), monocytes count ($*10^{9}$/L), platelets count ($*10^{9}$/L), and hemoglobin (g/dl);

**Blood biochemistry:** serum creatinine (umol/L), blood urea or blood urea nitrogen (mg/dl), and serum glucose (mmol/L);

**Coagulation function:** Prothrombin time (sec), activated partial thromboplastin time (sec), and international normalized ratio.

The laboratory test assessment will be collected at baseline and 24 ± 12 hours post treatment.

#### 2.2 Statistical Analysis Population

The analysis populations include intention-to-treat (ITT) population and per-protocol analysis set (PPS) for this study.

##### 2.2.1 Intention-to-treat Population

The ITT population will include all randomized subjects according to ITT principles, regardless of their eligibility and any protocol deviations, in which subjects will be analyzed according to the group assigned by randomization. ITT is the primary efficacy analysis set for this study and will also be used for analyses of demographics, baseline characteristics, medical history and concomitant medication information.

##### 2.2.2 Per-protocol Analysis Set

The PPS is a subset of ITT population, including all randomized subjects who have been treated in the study without major protocol deviations that may significantly impact the interpretation of efficacy results. Detailed protocol deviation criteria will be determined at the latest before database lock. Subjects entering PPS need to satisfy all the following basic criteria:

(1) Meet all the eligibility criteria specified in the study protocol;

(2) The subjects were randomized and received the assigned treatment.

(3) Have a blind assessment of the 90-day outcome.

#### 2.3 Statistical Methods

For continuous data, the following statistics will be provided: number, mean, standard deviation (SD), median, lower quartile (Q1), upper quartile (Q3), minimum and maximum, unless otherwise stated. Categorical data will be summarized in terms of the number of patients and percentages.

For summary statistics, mean, standard deviation, median and quartiles will be reported to one more decimal place than the original data, while the 95% confidence interval (CI) will be reported to 2 more decimal places. Minimum and maximum values will be reported to the same number of significant digits as the original data. In the frequency table, the percentages will keep one decimal and the p values will keep 4 decimal or will be displayed as "<0.0001".

##### 2.3.1 Subject disposition

The total number of screened subjects and the number and proportion of subjects by treatment groups who were randomized, received study treatment, completed the study, and stopped the study early, will be provided. The number and proportion of subjects according to reasons for withdrawal from the trial were summarized descriptively, and also summarized for each analysis population by treatment groups. Where necessary, the CONSORT flow chart will be presented to describe the study subject disposition in the statistical analysis report.

##### 2.3.2 Demography and baseline characteristics

Demographic data and other baseline characteristics will be summarized by treatment groups for this study, as follows:

- Age (years);
- Sex (female vs. male);
- Race (The Han Nationality, and Other);
- Weight (kg);
- Height (cm);
- Body mass index (BMI, $kg/m^2$) = weight (kg)/height $(m)^2$

In addition, smoking and drinking history, baseline mRS, baseline GCS, baseline NIHSS scale and other information will also be summarized by treatment groups. Data listings will be provided where necessary.

##### 2.3.3 Medical history

The medical history includes medical history of stroke, carotid artery disease, cardiac disorders, hypertension, diabetes mellitus, hypercholesterolaemia and allergy history et al.
Medical history will be summarized by treatment groups.

##### 2.3.4 Concomitant medication

Concomitant medication including thrombolytics, anticoagulants, antiplatelets, blood pressure/lipid and blood glucose control medications and other medications taken by the subjects during the study.

Concomitant medication will be summarized by treatment groups.

###### 2.3.5 Analysis of efficacy outcomes

All efficacy data analyses will be conducted based on ITT population; for primary and secondary outcome analysis, PPS will also be used as a supportive role.

###### 2.3.5.1 Primary efficacy outcome

The distribution of the mRS score at day 90 will be reported for each treatment group. The primary efficacy outcome analysis will be conducted by using ordinal logistic regression analysis adjusted for prognostic factors, including center as a random effect, preferred initial thrombectomy strategy (stent retriever vs. aspiration vs. stent retriever + aspiration), onset to randomization time (continuous), mRS before stroke (categorical), age (above vs. below median), and baseline NIHSS (above vs. below median) as fixed covariates. Adjusted and unadjusted common proportional odds ratio (cOR) for 1-point improvement in the mRS score will be derived from this model as treatment effect size (intervention group vs. control group) with their corresponding 95% confidence interval (CI). In case of deviations in proportional odds assumption, the mRS score at day 90 will be compared between groups by using the Mann-Whitney U test, and Wilcoxon-Mann-Whitney generalized OR will be calculated as effect size [2].

###### 2.3.5.2 Secondary efficacy outcomes

For binary efficacy outcomes (dichotomized mRS at day 90, reperfusion outcome in final angiogram, reperfusion outcome after the first pass, occurrence of emboli to a new territory, technical success rate and percentage of subjects with acceptable revascularization quality), between-group comparisons will be performed by using a Chi-squared test or Fisher's exact test when applicable. The binary secondary efficacy outcomes also will be analyzed by logistic regression analysis to provide an odds ratio and its 95% CI, if applicable. The adjustment factors are the same as those in the primary outcome analysis.

For continuous efficacy outcomes (infarction volume, the change in NIHSS score at 24 hours post treatment from baseline and change in NIHSS score at 7 days post treatment or discharge from baseline), between-group comparisons will be done using a linear model adjusted for the prognostic factors considered in the primary outcome analysis, as appropriate; the adjusted mean difference will be derived from a linear model as treatment effect sizes. In case of obvious deviation from normality of the model residuals (except if a logarithmic transformation or other common transformation could be applied), a non-parametric analysis with the Mann-Whitney U test will be used and standardized differences will be calculated. Rank-transformed data will be provided as treatment effect size.

For count variable (number of thrombectomy attempts), it will be analyzed by using a negative binomial model [3] with the count data as dependent variable and treatment group as fixed-effect factors adjusted for the prognostic factors considered in the primary outcome analysis, as appropriate. The model estimated event rates and its 95%

confidence intervals will be provided by treatment groups. The treatment comparison will be performed through the estimated ratio of risk rates.

For time-to-event variable (time from groin puncture to successful reperfusion), it will be analyzed using the Cox's proportional hazards model with a fixed treatment group factor and adjusted for the prognostic factors considered in the primary outcome analysis, as appropriate. The estimated hazard ratio and the corresponding two-sided 95% CI will be provided. The Kaplan-Meier curves by treatment groups will be presented.

###### 2.3.5.3 Subgroup analyses

For the primary outcome, pre-specified subgroup analyses will be performed by examining the interaction between specific baseline characteristics and treatment. Subgroups are defined as follows:

- age (above vs. below median)
- onset time to randomization (<6 vs. ≥6 h)
- NIHSS at baseline (above vs. below median)
- occlusion location (internal carotid artery vs. middle cerebral artery)
- presumed etiological AIS subtype (intracranial atherosclerosis vs. extracranial lesion vs. cardioembolism vs. other/undetermined)
- preferred treatment strategy (stent retriever vs. aspiration vs. stent retriever + aspiration)
- Tandem lesion (Yes vs. No)
- Intravenous thrombolysis (Yes vs. No)

The analysis for each subgroup will be performed by adding the subgroup variable as well as its interaction with the intervention as fixed effects to the main logistic regression model. Within each subgroup, summary measures will include raw counts and percentages by treatment arm, as well as the OR for treatment effect with a 95% CI. The output results will be displayed on a forest plot, including the p-values for heterogeneity corresponding to the interaction term between intervention and each subgroup variable.

##### 2.3.6 Safety analysis

In this study, the safety analysis will be mainly based on statistical description. All of the safety data analyses will be performed in both ITT and PPS.

###### 2.3.6.1 Analysis of AEs

The number and percentage of subjects who had at least one serious adverse event, classification of serious adverse event, adverse events of intracranial hemorrhage and symptomatic intracranial hemorrhage or asymptomatic intracranial hemorrhage at 7 days post treatment or discharge.

- All SAEs will be summarized by SOC and PT;
- Any peri-procedural complications will be summarized by SOC and PT.

In order to signal potential safety problems, comparisons of the frequency of SAEs and peri-procedural complications between treatment groups will be performed by using a Chi-squared test or Fisher's exact test when appropriate.

The data listings will be provided for All SAEs, any procedural complications and deaths at 90 days post treatment.

###### 2.3.6.2 Analysis of vital signs

Summaries of vital signs parameters will be presented by treatment groups, using summary statistics for observed values for each parameter.

###### 2.3.6.3 Clinical laboratory data analysis

For continuous laboratory parameters, the summary statistics will be provided by treatment groups for observed values for each parameter.

If a lab test result is recorded as "<10", then it will be summarized as a value of "5", if applicable; and likewise, ">10" will be summarized as "10".

##### 2.3.7 Cost outcomes analysis

For the EQ-5D dimensions, between-group comparisons will be done using a linear model adjusted for the prognostic factors considered in the randomization, as appropriate; the adjusted mean difference will be derived from a linear model as treatment effect sizes.

For other cost outcomes, they will need time to acquire and analyze, and hence will be presented in a secondary analysis and publication, which is out of the analysis scope of the SAP.

#### 2.4 Data Processing Conventions

##### 2.4.1 Baseline definition

In this study, baseline values are defined as those data collected before intervention (e.g. at baseline visit). When multiple data collections occur during the baseline period, the final data shall prevail in principle, unless explicitly stated.

##### 2.4.2 Missing data

We will report proportions of missing values for all collected variables where needed.

Missing baseline characteristics data will be imputed by regression interpolation as appropriate.

If there is a large number of missing data on efficacy and safety, an evaluation on the missing data should be conducted before formal analysis, and the study team will propose and determine the solution before database lock.

For subjects who died within the study period, the worst scores will be assigned for all subsequent not-assessed clinical outcome measures in their analyses, as follows (Table 3).

**Table 3. The worst scores of clinical outcomes**

| Clinical outcomes | The worst scores |
|---|---|
| mRS | 6 |
| NIHSS | 42 |

##### 2.4.3 Time window

Not applicable.

##### 2.4.4 Unscheduled visits

Not applicable.

##### 2.4.5 Centers pooling

This study will not pool and analyze the data of each study center, except including center as a random effect in the analyses of efficacy outcomes.

### 3 CHANGES TO PLANNED ANALYSES FROM THE PROTOCOL

No changes of planned analyses in the protocol are made in this statistical analysis plan.

### 4 INTERIM ANALYSIS

The study consists of 2 interim analyses to be performed when 30% and 60% of the 90-day follow-up data have been collected.

The independent data and safety monitoring board (DSMB) will adopt the Haybittle-Peto rule, and $\alpha$ <0.001 in the interim analysis will be considered as statistically significant. Because 2 interim analyses will be performed as scheduled, the significance level of $\alpha = 0.0482$ will be used for final analysis.

The DSMB will monitor SAEs (e.g. death, spontaneous intracerebral hemorrhage, and neurological deterioration) periodically, and the occurrence of excessive SAEs will trigger discussion regarding termination of the study.

### 5 STATISTICAL ANALYSIS SOFTWARE

All statistical analyses and data summary will be carried out by using SAS® 9.4 in this study. Software R 4.0.1 or higher version will be used for drawing plots if applicable.

### 7 APPENDIX

Appendix 1: NIHSS Scale

The NIHSS is an ordinal hierarchical scale that evaluates the severity of stroke by assessing the patient's physical strength. Scores may range from 0 to 42, with higher scores indicating more severe ischemia. Administer stroke scale items in the order listed. Record performance in each category after each subscale exam. Do not go back and change scores. Follow directions provided for each exam technique. Scores should reflect what the patient does, not what the clinician thinks the patient can do. The clinician should record answers while administering the exam and work quickly. Except where indicated, the patient should not be coached (i.e., repeated requests to patient to make a special effort).

| Instructions | Scale Definition |
|---|---|
| **1a. Level of Consciousness.** The investigator must choose a response if a full evaluation is prevented by such obstacles as an endotracheal tube, language barrier, orotracheal trauma/bandages. A 3 is scored only if the patient makes no movement (other than reflexive posturing) in response to noxious stimulation. | **0=Alert**; keenly responsive.<br>**1=Not alert**; but arousable by minor stimulation to obey, answer, or respond.<br>2=**Not alert**; requires repeated stimulation to attend, or is obtunded and requires strong or painful stimulation to make movements (not stereotyped).<br>3=Responds only with reflex motor or autonomic effects, or totally unresponsive, flaccid, and areflexic. |
| **1b. Level of Consciousness Questions:** The patient is asked the month and his/her age. The answer must be correct - there is no partial credit for being close. Aphasic and stuporous patients who do not comprehend the questions will score 2. Patients unable to speak because of endotracheal intubation, orotracheal trauma, severe dysarthria from any cause, language barrier, or any other problem not secondary to aphasia are given a 1. It is important that only the initial answer be graded and that the examiner not "help" the patient with verbal or non-verbal cues. | 0=Answers both questions correctly.<br>1=Answers one question correctly.<br>2=Answers neither question correctly. |
| **1c. LOC Commands:** The patient is asked to open and close the eyes and then to grip and release the non-paretic hand. Substitute another one-step command if the hands cannot be used. Credit is given if an unequivocal attempt is made but not completed due to weakness. If the patient does not respond to command, the task should be demonstrated to him or her (pantomime), and the result scored (i.e., follows none, one, or two commands). Patients with trauma, amputation, or other physical impediments should be given suitable one-step commands. Only the first attempt is scored. | 0=Performs both tasks correctly.<br>1=Performs one task correctly.<br>2=Performs neither task correctly. |
| **2. Best Gaze:** Only horizontal eye movements | **0=Normal.** |

| Instructions | Scale Definition |
|---|---|
| will be tested. Voluntary or reflexive (oculocephalic) eye movements will be scored, but caloric testing is not done. If the patient has a conjugate deviation of the eyes that can be overcome by voluntary or reflexive activity, the score will be 1. If a patient has an isolated peripheral nerve paresis (CN III, IV, or VI), score a 1. Gaze is testable in all aphasic patients. Patients with ocular trauma, bandages, pre-existing blindness, or other disorder of visual acuity or fields should be tested with reflexive movements, and a choice made by the investigator. Establishing eye contact and then moving about the patient from side to side will occasionally clarify the presence of a partial gaze palsy. | **1=Partial gaze palsy;** gaze is abnormal in one or both eyes, but forced deviation or total gaze paresis is not present.<br>2=**Forced deviation,** or total gaze paresis is not overcome by the oculocephalic maneuver. |
| **3. Visual:** Visual fields (upper and lower quadrants) are tested by confrontation, using finger counting or visual threat, as appropriate. Patients may be encouraged, but if they look at the side of the moving fingers appropriately, this can be scored as normal. If there is unilateral blindness or enucleation, visual fields in the remaining eye are scored. Score 1 only if a clear-cut asymmetry, including quadrantanopia, is found. If patient is blind from any cause, score 3. Double simultaneous stimulation is performed at this point. If there is extinction, patient receives a 1, and the results are used to respond to item 11. | **0=No visual loss.**<br>**1=Partial hemianopia.**<br>**2=Complete hemianopia.**<br>**3=Bilateral hemianopia** (blind including cortical blindness). |
| **4. Facial Palsy:** Ask or use pantomime to encourage the patient to show teeth or raise eyebrows and close eyes. Score symmetry of grimace in response to noxious stimuli in the poorly responsive or non-comprehending patient. If facial trauma/bandages, orotracheal tube, tape, or other physical barriers obscure the face, these should be removed to the extent possible. | **0=Normal symmetrical movements.**<br>**1=Minor paralysis** (flattened nasolabial fold, asymmetry on smiling).<br>**2=Partial paralysis** (total or near-total paralysis of lower face).<br>**3=Complete paralysis of one or both sides** (absence of facial movement in the upper and lower face). |
| **5. Motor Arm:** The limb is placed in the appropriate position: extend the arms (palms down) 90 degrees (if sitting) or 45 degrees (if supine). Drift is scored if the arm falls before 10 seconds. The aphasic patient is encouraged using urgency in the voice and pantomime, but not noxious stimulation. Each limb is tested in turn, beginning with the non-paretic arm. Only in the case of amputation or joint fusion at the shoulder, the examiner should record the score as untestable (UN) and clearly write the explanation for this choice. | **0=No drift**; limb holds 90 (or 45) degrees for full 10 seconds.<br>**1=Drift;** limb holds 90 (or 45) degrees, but drifts down before full 10 seconds; does not hit bed or other support.<br>**2=Some effort against gravity;** limb cannot get to or maintain (if cued) 90 (or 45) degrees, drifts down to bed, but has some effort against gravity.<br>**3=No effort against gravity**; limb falls.<br>**4=No movement.**<br>UN=Amputation or joint fusion: explain:<br>**5a=left arm.** |

| Instructions | Scale Definition |
|---|---|
| | **5b=right arm.** |
| **6. Motor Leg:** The limb is placed in the appropriate position: hold the leg at 30 degrees (always tested supine). Drift is scored if the leg falls before 5 seconds. The aphasic patient is encouraged using urgency in the voice and pantomime, but not noxious stimulation. Each limb is tested in turn, beginning with the non-paretic leg. Only in the case of amputation or joint fusion at the hip, the examiner should record the score as untestable (UN) and clearly write the explanation for this choice. | **0=No drift;** leg holds 30-degree position for full 5 seconds.<br>**1=Drift;** leg falls by the end of the 5-second period but does not hit the bed or other support.<br>**2=Some effort against gravity**; leg falls to bed by 5 seconds but has some effort against gravity.<br>**3=No effort against gravity**; leg falls to bed immediately.<br>**4=No movement.**<br>UN=Amputation or joint fusion: explain:<br>**6a. Left leg**<br>**6b. Right leg.** |
| **7. Limb Ataxia:** This item is aimed at finding evidence of a unilateral cerebellar lesion. Test with eyes open. In case of visual defect, ensure testing is done in intact visual field. The finger-nose-finger and heel-shin tests are performed on both sides, and ataxia is scored only if present out of proportion to weakness. Ataxia is absent in the patient who cannot understand or is paralyzed. Only in the case of amputation or joint fusion, the examiner should record the score as untestable (UN) and clearly write the explanation for this choice. In case of blindness, test by having the patient touch nose from extended arm position. | **0= Absent.**<br>**1=Present in one limb.**<br>**2=Present in two limbs.**<br>UN=Amputation or joint fusion: explain: |
| **8. Sensory:** Sensation or grimace to pinprick when tested, or withdrawal from noxious stimulus in the obtunded or aphasic patient. Only sensory loss attributed to stroke is scored as abnormal and the examiner should test as many body areas [arms (not hands), legs, trunk, face] as needed to accurately check for hemisensory loss. A score of 2, "severe or total sensory loss," should only be given when a severe or total loss of sensation can be clearly demonstrated. Stuporous and aphasic patients will, therefore, probably score 1 or 0. The patient with brainstem stroke who has bilateral loss of sensation is scored 2. If the patient does not respond and is quadriplegic, score 2. Patients in a coma (item 1a=3) are automatically given a 2 on this item. | **0=Normal;** no sensory loss.<br>**1=Mild-to-moderate sensory loss;** patient feels pinprick is less sharp or is dull on the affected side; or there is a loss of superficial pain with pinprick, but patient is aware of being touched.<br>**2=Severe or total sensory loss;** patient is not aware of being touched in the face, arm, and leg. |
| **9. Best Language:** A great deal of information about comprehension will be obtained during the preceding sections of the examination. For this scale item, the patient is asked to describe what is happening in the attached picture, to name the items on the attached naming sheet, and to read | **0=No aphasia;** normal.<br>**1=Mild-to-moderate aphasia;** some obvious loss of fluency or facility of comprehension, without significant limitation on ideas expressed or form of expression. Reduction of speech and/or |

| Instructions | Scale Definition |
|---|---|
| from the attached list of sentences. Comprehension is judged from responses here, as well as to all of the commands in the preceding general neurological exam. If visual loss interferes with the tests, ask the patient to identify objects placed in the hand, repeat, and produce speech. The intubated patient should be asked to write. The patient in a coma (item 1a=3) will automatically score 3 on this item. The examiner must choose a score for the patient with stupor or limited cooperation, but a score of 3 should be used only if the patient is mute and follows no one-step commands. | comprehension, however, makes conversation about provided materials difficult or impossible. For example, in conversation about provided materials, examiner can identify picture or naming card content from patient's response.<br>**2=Severe aphasia;** all communication is through fragmentary expression; great need for inference, questioning, and guessing by the listener. Range of information that can be exchanged is limited; listener carries burden of communication. Examiner cannot identify materials provided from patient response.<br>**3=Mute, global aphasia;** no usable speech or auditory comprehension. |
| **10. Dysarthria:** If patient is thought to be normal, an adequate sample of speech must be obtained by asking patient to read or repeat words from the list in the attachment. If the patient has severe aphasia, the clarity of articulation of spontaneous speech can be rated. Only if the patient is intubated or has other physical barriers to producing speech, the examiner should record the score as untestable (UN) and clearly write the explanation for this choice. Do not tell the patient why he/she is being tested. | **0=Normal.**<br>**1=Mild-to-moderate dysarthria**; patient slurs at least some words and, at worst, can be understood with some difficulty.<br>**2=Severe dysarthria:** patient's speech is so slurred as to be unintelligible in the absence of or out of proportion to any dysphasia, or is mute/anarthric.<br>UN=Intubated or other physical barrier. |
| **11. Extinction and Inattention (formerly Neglect):** Sufficient information to identify neglect may be obtained during the prior testing. If the patient has a severe visual loss preventing visual double simultaneous stimulation, and the cutaneous stimuli are normal, the score is normal. If the patient has aphasia but does appear to attend to both sides, the score is normal. The presence of visual spatial neglect or anosagnosia may also be taken as evidence of abnormality. Since the abnormality is scored only if present, the item is never untestable. | **0=No abnormality.**<br>**1=Visual, tactile, auditory, spatial, or personal inattention,** or extinction to bilateral simultaneous stimulation in one of the sensory modalities.<br>**2=Profound hemi-inattention or extinction to more than one modality;** does not recognize own hand or orients to only one side of space. |

#### Appendix 2: eTICI Scale

| Grade | Short description | Long description |
|---|---|---|
| 0 | No perfusion | No antegrade flow beyond the point of occlusion. |
| 1 | Minimal perfusion | The antegrade flow passes the area of obstruction but there is little or slow reperfusion flow in the distal branches. |
| 2a | Reperfusion <50% | Antegrade flow fills less than 50% of the ischemic areas of target vessel occlusion (e.g., 1 important branch of the middle cerebral artery and its territory). |
| 2b | 50% ≤ Reperfusion <90% | Antegrade flow fills more than 50% of the ischemic areas of target vessel occlusion (e.g., 2 important branches of the middle cerebral artery and their territories). |
| 2c | Reperfusion ≥90% | Reperfusion antegrade flow almost completely fill the whole ischemic areas of the target vessel occlusion, with only slow flow or thrombus in a small portion of the distal vessel branches. |
| 3 | 100% reperfusion | Reperfusion antegrade flow completely fill the whole ischemic areas of the target vessel occlusion, with no visible occlusion in the distal branches. |

#### Appendix 3: EQ-5D Health-Related Quality of Life Scale

| Date of score: \|__\|__\|/\|__\|__\|/\|__\|__\|__\|__\| (DD/MM/YYYY) | |
|---|---|
| **Item** | **Score** |
| **Mobility** | □ I have no problems in walking about<br>□ I have slight problems in walking about<br>□ I have moderate problems in walking about<br>□ I have severe problems in walking about<br>□ I am unable to walk about |
| **Self-care** | □ I have no problems washing or dressing myself<br>□ I have slight problems washing or dressing myself<br>□ I have moderate problems washing or dressing myself<br>□ I have severe problems washing or dressing myself<br>□ I am unable to wash or dress myself |
| **Usual activities** | □ I have no problems doing my usual activities<br>□ I have slight problems doing my usual activities<br>□ I have moderate problems doing my usual activities<br>□ I have severe problems doing my usual activities<br>□ I am unable to do my usual activities |
| **Pain/Discomfort** | □ I have no pain or discomfort<br>□ I have slight pain or discomfort<br>□ I have moderate pain or discomfort<br>□ I have severe pain or discomfort<br>□ I have extreme pain or discomfort |
| **Anxiety/Depression** | □ I am not anxious or depressed<br>□ I am slightly anxious or depressed<br>□ I am moderately anxious or depressed<br>□ I am severely anxious or depressed<br>□ I am severely anxious or depressed |
| **Status that best describes your health** | Score _________ (0-100) |
