## Supplementary material for "Proxiaml Temporary Occlusion using Balloon Guide Catheter for Mechanical Thrombectomy (PROTECT-MT): Study protocol for a multicenter randomized trial": protocol version 4.0

Project Leader: Professor Jianmin Liu

Contract Research Organization: CCRC Medtech (Shanghai) Co., Ltd.

Protocol No.: PROTECT-MT

Protocol Version: V4.0

Protocol Date: Oct 25, 2023

11.3 Approval of the Study Protocol ... 29
**12 Quality Control of the Study ... 29**
12.1 Investigators Training ... 29
12.2 Measures to Improve Subjects Compliance ... 29
12.3 Clinical Monitoring ... 30
12.4 Data Management ... 30
12.5 Maintenance of Source Documents ... 30
**13 Study Administration ... 30**
13.1 Organizational Structure ... 30
13.1.1 Steering Committee ... 31
13.1.2 Central Coordinating Center (CCC) ... 31
13.1.3 Imaging Adjudication Committee (Core Laboratory) ... 31
13.1.4 Clinical Events Committee (CEC) ... 31
13.1.5 Data and Safety Monitoring Board (DSMB) ... 32
13.1.6 Outcome Assessment Committee (OAC) ... 32
13.2 List of Study Sites ... 32
13.3 Confidentiality Principle ... 32
13.4 Agreement on the Publication of Study Results ... 33
**14 Fund ... 33**
**15 Appendix ... 35**
**16 References ... 42**

### Protocol Signature Page

#### Signature of Sponsor

Study Title: PROximal TEmporary oCclusion using balloon guide caTheter for Mechanical Thrombectomy (PROTECT-MT)

Protocol No.: PROTECT-MT

Version No.: V4.0

Version Date: Oct 25, 2023

| Name of Sponsor | Signature of Project Leader | Date |
|---|---|---|
| The First hospital Affiliated to Naval Medical University | | |

I have carefully read the contents of this clinical study protocol and confirm that all relevant contents are included in it. I will conduct this clinical study in strict compliance with the Declaration of Helsinki, current laws and regulations in China, as well as this clinical study protocol.

I will be responsible for providing this clinical study protocol and relevant information to team members involved in this study and fully discuss it with them to ensure that they completely understand the contents of this study protocol and the study procedures.

I will be responsible for accurately recording all required data in the Case Report Form (CRF) and cooperate in completing the clinical study report. I agree to allow the monitors, auditors and regulatory authorities to monitor, audit and inspect this clinical study.

| Name of the Principal Investigator | Signature | Date |
|---|---|---|
| Jianmin Liu | | |

### Signature of Site

Study Title: PROximal TEmporary oCclusion using balloon guide caTheter for Mechanical Thrombectomy (PROTECT-MT)

Protocol No.: PROTECT-MT

Version No.: V4.0

Version Date: Oct 25, 2023

I have carefully read the contents of this clinical study protocol and confirm that all relevant contents are included in it. I will conduct this clinical study in strict compliance with the Declaration of Helsinki, current laws and regulations in China, as well as this clinical study protocol.

I will be responsible for providing this clinical study protocol and relevant information to team members involved in this study and fully discuss it with them to ensure that they completely understand the contents of this study protocol and the study procedures.

I will be responsible for accurately recording all required data in the Case Report Form (CRF) and cooperate in completing the clinical study report. I agree to allow the monitors, auditors and regulatory authorities to monitor, audit and inspect this clinical study.

| **Name of the Principal Investigator** | **Signature** | **Date** |
|---|---|---|

#### List of Abbreviations

| Abbreviation | Definition |
|---|---|
| AE | Adverse Event |
| AIS | Acute Ischemic Stroke |
| CRA | Clinical Research Associate |
| CRO | Contract Research Organization |
| CRF | Case Report Form |
| CT | Computed Tomography |
| CTA | Computed Tomography Angiography |
| DSA | Digital Substraction Angiography |
| EC | Ethics Committee |
| EDC | Electric Data Capture |
| eTICI | Extended Thrombolysis in Cerebral Infarction |
| GCS | Glasgow Coma Scale |
| ICF | Informed Consent Form |
| MRA | Magnetic Resonance Angiography |
| MRI | Magnetic Resonance Imaging |
| mRS | Modified Rankin Scale |
| NIHSS | National Institute of Health Stroke Scale |
| PI | Principal Investigator |
| SAE | Serious Adverse Event |

**Protocol Synopsis**

| | |
|---|---|
| **Sponsor** | The First hospital Affiliated to Naval Medical University(Changhai hospital) |
| **Protocol No.** | PROTECT-MT |
| **Study Title** | PROximal TEmporary oCclusion using balloon guide caTheter for Mechanical Thrombectomy (PROTECT-MT) |
| **Study Design** | A prospective, multicenter, randomized controlled, open-label, blinded outcome evaluation (PROBE) trial |
| **Study Population** | Subjects with acute ischemic stroke(AIS) due to anterior circulation large vessel occlusion(LVO), and are eligible for mechanical thrombectomy(MT) according to local guidelines |
| **Study Objective** | To determine the effectiveness of balloon guide catheter(BGC) as compared to standard guide catheter on functional outcome (modified Rankin Scale [mRS] score) in patients with acute ischemic stroke due to anterior circulation large vessel occlusion. |
| **Number of planed Sites** | 40-60 centers |
| **Inclusion/Exclusion Criteria** | **Inclusion Criteria**<br>To be eligible for inclusion for this study, subjects are to satisfy all of the following criteria:<br>1) Age ≥18 years.<br>2) Diagnosis of AIS with confirmed anterior circulation LVO (including intracranial segment of the internal carotid artery, and the first or proximal second segment [M1 or proximal M2] of the middle cerebral artery) by brain imaging.<br>3) To receive MT within 24 hours after AIS onset according to local guidelines.<br>4) Preoperative mRS score of 0-1.<br>5) Signed informed consent form obtained from the subject (or approved surrogate).<br>**Exclusion Criteria**<br>Subjects who meet any of the following criteria should be excluded from this study:<br>1) Intracranial hemorrhage confirmed by imaging.<br>2) Known or suspected pre-existing (chronic) large vessel occlusion in the symptomatic territory.<br>3) Excessive vascular access tortuosity disables the use of balloon guide catheter.<br>4) Intracranial stent implanted in the symptomatic territory that precludes the deployment/removal of the mechanical thrombectomy device.<br>5) Any other condition that precludes the performing of mechanical thrombectomy procedure. |

| | |
|---|---|
| | 6) Occlusions in multiple vascular territories confirmed by Computed Tomography Angiography(CTA) or Magnetic Resonance Angiography(MRA).<br>7) Subjects who are pregnant.<br>8) Subjects who are allergy to the contrast agent.<br>9) Subjects who refuse to cooperate or unable to tolerate interventional operation.<br>10) Subjects whose expected lifetime are less than 90 days.<br>11) Subjects who are unlikely to participate in follow-up assessments according to the investigator's judgement.<br>12) Any other condition that, according to the investigator's judgement, not suitable for using of balloon guide catheter. |
| **Randomization, Group Assignment, and Blinding** | 1. **Randomization:**<br>After confirmation of eligibility, subjects will be randomized via an Internet-based randomization system in a 1:1 manner to treatment with BGCs(Intervention group) or not(control group), stratified by site, preferred thrombectomy strategy(stent retriever VS aspiration VS stent retriever + aspiration), and onset to randomization time( <6 hours VS ≥6 hours)<br>**2. Group Assignment:**<br>Subjects will be treated with thrombectomy based on the results of randomization.<br>**Intervention group:** Subjects treated with BGCs combined with conventional thrombectomy.<br>**Control group:** Subjects treated with standard guide catheter combined with conventional thrombectomy.<br>Any National Medical Products Administration (NMPA) devices (including the use of a thrombectomy stent, distal aspiration catheter, or combine use of both) are allowed.<br>**3. Blinding:**<br>Both patient and treating physician will be aware of the treatment assignment. But outcome evaluation would be blinded. Information on outcome at 90 days will be assessed through standardized forms and procedures, by a trained investigator blinded for treatment allocation. Interviews will be recorded. Assessors who are blinded to the treatment allocation will perform assessment of outcome on the modified Rankin scale on this information. Results of neuro-imaging will be also assessed in a blinded manner. Information on treatment allocation will be kept separate from the main study database. The steering committee will be kept unaware of the results of interim analyses of efficacy and safety. An independent DSMB statistician will combine data on treatment allocation with the clinical data in order to report to the data monitoring committee (DSMB). |
| **Study Outcomes** | **1. Primary Outcomes**<br>Functional outcome, defined as modified Rankin Scale (mRS) score shift at 90 days (±14 days) .<br>**2. Secondary Outcomes** |

| | |
|---|---|
| | 3%; mRS 1: 9%; mRS 2: 21%; mRS 3: 18%; mRS 4: 22%; mRS 5: 6% and mRS 6:21%. Based on previous studies, we assumed a favorable treatment effect with a common odds ratio (cOR) of 1.43, corresponding to an 8% absolute increase in the rate of mRS scores of 0-2. In a simulation with 5000 runs we computed the proportion of positive trials, for a given sample size. A sample size of 920 was determined to detect the superiority with a power of 87% and two-sided alpha of 0.05. Considering 5% dropout rate and 5% crossover rate, a final sample size of 1074, 537 per arm, would be needed.<br>• **Methods for Statistical Analysis**<br>Analyses will be performed using the intention-to-treat (ITT) principle.<br>Statistical analysis will be detailed in a pre-specified Statistical Analysis Plan (SAP).<br>Two interim analyses would be performed when 30% and 60% of the 90-day follow-up data have been collected. |

### 1 Study Background

Stroke has become the leading cause of death and adult disability in China [1]. According to Stroke Prevention and Treatment Report of China in 2019, there are 12.42 million stroke cases in people aged 40 years or above in China, 65% of whom are ischemic stroke. With population aging, stroke prevalence is continuously rising, bringing a heavy burden to the families and society [2].

The key to the treatment of acute ischemic stroke (AIS) is to recanalize the occluded vessels as early as possible and save the brain tissue around the core infarct area, namely the ischemic penumbra. With the publication of five randomized controlled trials comparing Endovascular treatment (EVT) with standard medical treatment in 2015 [3-7], EVT combined with intravenous thrombolysis has become the preferred treatment for patients with large vessel occlusion of anterior circulation within 6 hours of onset [8]. Subsequently, the DAWN and DEFUSE 3 studies in 2018 further extended the treatment time window to 24 hours [9, 10]. Although mechanical thrombectomy has significantly improved the successful recanalization rate in patients with acute ischemic stroke due to large vessel occlusion, a large proportion of patients still have poor clinical prognosis. How to improve the prognosis of these patients remain a clinical problem needed to be solved.

The Balloon Guide Catheter(BGC) is device used during mechanical thrombectomy, which offers proximal flow control and a stable platform to facilitate the insertion and guidance of an intravascular catheter. Dilation of the balloon during the procedure would enable a significant flow arrest or reversal, and hence reduce the risk of thrombus fragmentation and distal embolism [11]. Some observational studies [12-17] have shown that the use of balloon guide catheter could improve the quality of final reperfusion, shorten the procedure time, and may lead to better clinical outcomes in patients undergoing thrombectomy. In a retrospective study involving 955 patients, the final and first-pass reperfusion rates (mTICI 2b or 3) were 86.8% and 37.0% in BGC group, which were higher than those in the standard guide catheter group,i.e 74.7% and 14.1% ($P<0.001$). Thrombectomy maneuvers (2.5±1.9 versus 3.3±2.1; $P<0.001$) and procedure time (54.3±27.4 min vs 67.6±38.2 min; $P<0.001$)were significantly decreased in BGC group compared with those in standard guide catheter group. In multivariate analysis, BGC usage was an independent factor for good clinical outcome (odds ratio: 1.40; 95% CI, 1.02 1.92; $P = 0.038$) [16]. In another prospective registry study, STRATIS (Systematic Evaluation of Patients Treated With Neurothrombectomy Devices for Acute Prevention Stroke), the first recertification (≥2c and ≥2b) rates of three guide catheter(BGC, standard guide catheter, distal access catheter ) were (212/443 [48%] vs 16/62 [26%] vs 83/235 [35%]), and (294/443 [66%] vs 26/62 [42%] vs 129/234 [55%]), respectively. The first recanalization rate of BGC group was higher than that of distal access catheter and standard guide catheter, and the clinical prognosis of patients in the balloon guide catheter group was also better than that of the other two groups, and the rates of functional independence were 253/415 [61%] vs. 23/55 [42%] vs. 113/218 [52%] [14].

However, controversies existed regarding the use of balloon guide catheter. For example, Bourcier R et al reported that balloon-guide catheter may be unnecessary with used with a combination of aspiration catheter and stent, and the rates of recanalization (60.1% VS 62.7%; RR, 0.92; 95%CI, 0.80-1.14) and functional outcome (44.6% Vs 40.0%; RR 1.12, 95%CI, 0.85-1.47) were similar between BGC group and standard guide catheter group [18]. In addition, some technical issues may hinder the widespread use of balloon guide catheters. The stiffness and trackability of the catheter may result in complications, also preparing and delivering the BGCs may prolonged procedure time and increase of the cost of the procedure [19]. These issues have aroused debates on whether balloon guide catheter should be routinely used in thrombectomy treatment, which led to absence of standardized procedures for thrombectomy[20, 21].

Because of the above reasons, there is an urgent need for a high-quality randomized trial to assess the effectiveness of BGC in thrombectomy for acute stroke due to large vessel occlusion and to standardize the procedure of mechanical thrombectomy. We aimed at establishing the best thrombectomy technique and procedure for AIS due to LVO, so as to improve the clinical prognosis, and reduce the national economy burden.

#### 2 Study Objective

The objective of this study is to determine the effectiveness of balloon guide catheter(BGC) as compared to standard guide catheter on functional outcome (modified Rankin Scale [mRS] score at Day 90) in patients with acute ischemic stroke due to anterior circulation large vessel occlusion.

#### 3 Study Design

This is a prospective, multicenter, randomized controlled, open-label, blinded outcome assessment trial involving 1074 eligible subjects to be recruited from 40-60 hospitals in China.

Subjects who meet the eligibility criteria will be randomized to either intervention group (receiving treatment with BGC) or control group (receiving treatment with standard guide catheter). Followed up would be conducted at 1 day, 7 days, and 90 days after thrombectomy.

Study design is presented as below (Figure 1).

**Figure 1. Study schema**

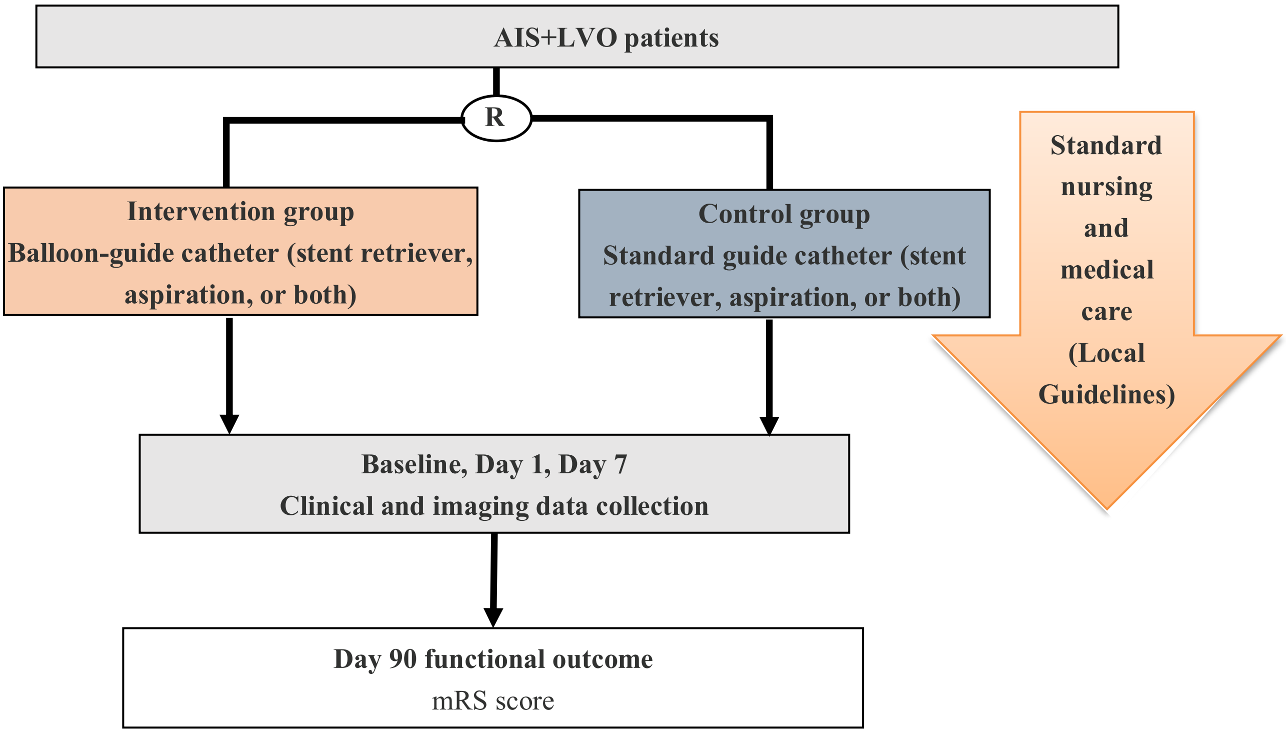

#### 4 Study Population

Subjects who are with AIS due to LVO of the anterior circulation confirmed by cranial imaging and judged to be eligible for being treated with mechanical thrombectomy according to local guidelines will be enrolled into this study. The principal investigator at each site will be responsible for subject screening and enrollment.

##### 4.1 Inclusion Criteria

To be eligible for inclusion for this study, subjects are to satisfy all of the following criteria:

1) Age ⩾18 years.
2) Diagnosis of AIS with confirmed anterior circulation LVO (including intracranial segment of the internal carotid artery, and the first or proximal second segment [M1 or proximal M2] of the middle cerebral artery) by brain imaging.
3) To receive MT within 24 hours after AIS onset according to local guidelines.
4) Preoperative mRS score of 0-1.

5) Signed informed consent form obtained from the subject (or approved surrogate).

#### 4.2 Exclusion Criteria

Subjects who meet any of the following criteria should be excluded from this study:

1) Intracranial hemorrhage confirmed by imaging.
2) Known or suspected pre-existing (chronic) large vessel occlusion in the symptomatic territory.
3) Excessive vascular access tortuosity disables the use of balloon guide catheter.
4) Intracranial stent implanted in the symptomatic territory that precludes the deployment/removal of the mechanical thrombectomy device.
5) Any other condition that precludes the performing of mechanical thrombectomy procedure.
6) Occlusions in multiple vascular territories confirmed by Computed Tomography Angiography(CTA) or Magnetic Resonance Angiography(MRA).
7) Subjects who are pregnant.
8) Subjects who are allergy to the contrast agent.
9) Subjects who refuse to cooperate or unable to tolerate interventional operation.
10) Subjects whose expected lifetime are less than 90 days.
11) Subjects who are unlikely to participate in follow-up assessments according to the investigator's judgement.
12) Any other condition that, according to the investigator's judgement, not suitable for using of balloon guide catheter.

#### 4.3 Subject Withdrawal Criteria

While study withdrawal is discouraged, subjects may withdraw from the study at any time, with or without reason and without prejudice to further treatment. Withdrawn subjects will not undergo any additional follow-up, nor will they be replaced, but the collected data may be used for statistical analysis. Such withdrawal, including the reason for withdrawal, should be documented in the patient's file.

#### 4.4 Premature Termination of the Study

This study could be terminated prematurely by the sponsor (i.e.due to finance or administrative reasons et al), regulatory authority, or the ethics committees from the perspective of protecting the subjects' rights and interests. In case of premature termination of the study, no subjects would be enrolled, the database will be closed after 90 days assessment of the last enrolled patient and results will be reported.

### 5 Randomization and Treatment allocation

Subjects will be randomized via an Internet-based randomization system in a 1:1 manner to treatment with BGCs(Intervention group) or not(control group), stratified by site, preferred initial thrombectomy strategy(stent retriever VS aspiration VS stent retriever + aspiration), and onset to randomization time( <6 hours VS ≥6 hours). The randomization sequence will use a minimization algorithm to ensure balance in key prognostic factors. Randomization is allowed when large vessel occlusion has been established.

Subjects will be treated with thrombectomy based on the results of randomization.

**Intervention group**: Subjects treated with BGCs combined with conventional thrombectomy.

**Control group**: Subjects treated with standard guide catheter combined with conventional thrombectomy.

Any National Medical Products Administration (NMPA) devices (including the use of a thrombectomy stent, distal aspiration catheter, or combine use of both) are allowed.

### 6 Study Method

#### 6.1 Study Outcomes

##### 6.1.1 Primary Outcomes

The primary outcome is functional outcome, defined as modified Rankin Scale (mRS) score shift at 90 days (±14 days).

The modified Rankin Scale (mRS) is an ordinal hierarchical scale ranging from 0 to 5, with higher scores indicating more severe disability. A score of 6 has been added to signify death.:

##### 6.1.4 Cost Outcomes

1) Health-related quality of life using EQ-5D (see **Appendix 3**);
2) Utility-weighted mRS scores;
3) Duration of hospitalization;
4) Treatment cost.

##### 6.1.5 Blinding

Both patient and treating physician will be aware of the treatment assignment. But outcome evaluation would be blinded. Information on outcome at 90 days will be assessed through standardized forms and procedures, by a trained investigator blinded for treatment allocation. Interviews will be recorded. Assessors who are blinded to the treatment allocation will perform assessment of outcome on the modified Rankin scale on this information. Results of neuro-imaging will be also assessed in a blinded manner**.** Information on treatment allocation will be kept separate from the main study database. The steering committee will be kept unaware of the results of interim analyses of efficacy and safety. An independent DSMB statistician will combine data on treatment allocation with the clinical data in order to report to the data monitoring committee (DSMB).

#### 6.2 Study Procedures

##### 6.2.1 Schedule of Study Activities

The study procedure is shown in the table below.

| Procedure/Time Window | Baseline | 1 day post treatment | 7 days post treatment# | 90 days post treatment |
|---|---|---|---|---|
| | Pre-operation - the day of treatment* | ±12 hours | ±2 days | ±14 days |
| Informed consent | X | | | |
| Demographics [1] | X | | | |
| Medical history [2] | X | | | |
| Vital signs [3] | X | X | X | |
| Blood routine test [4] | X | X | | |
| Blood biochemistry [5] | X | X | | |
| Coagulation function test [6] | X | X | | |
| Imaging examination [7] | X | X | X | |
| mRS score[8] | X | | | X |
| GCS score [9] | X | X | X | |
| NIHSS score [10] | X | X | X | |
| Quality of life using EQ-5D [11] | | | | X |
| Inclusion/exclusion criteria evaluation [12] | X | | | |
| Randomization and thrombectomy treatment [13] | X | | | |
| Medical cost | | | X | X |
| Safety evaluation [14] | X | X | X | X |
| Concomitant medication [15] | X | X | X | X |

Notes: #: at discharge or 7 days post treatment, whichever occurs first. *: The end date of surgery will be considered as the day of surgery.

1. **Demographics:** Age, sex, height and weight;
2. **Medical history:** Including medical history of stroke, carotid artery disease, cardiac disorders, peripheral arterial disease, hypertension, diabetes mellitus, and hypercholesterolaemia et al, stroke risk factors including history of smoking, alcohol drinking etc, and history of allergy (allergen);
3. **Vital signs:** Pulse, blood pressure, respiration rate, and body temperature;
4. **Blood routine:** Red blood cells (RBC) count, white blood cells (WBC) count, neutrophils count, lymphocytes count, monocytes count, platelets (PLT) count, and hemoglobin (HGB);
5. **Blood biochemistry:** Creatinine (Cr), blood urea or blood urea nitrogen (Urea/BUN), and glucose (Glu);
6. **Coagulation function:** Prothrombin time (PT), activated partial thromboplastin time (APTT), and international normalized ratio (INR);

7. **Imaging examination:** Data of brain imaging examinations (such as CT/CTA, MRI/MRA or DSA) performed at each visit will be collected;
8. **mRS score:** mRS score of subjects before this onset and mRS score at 90 days post treatment will be collected;
9. **GCS score:** GCS scores immediately before randomization, and at 1 day and 7 days post treatment will be collected;
10. **NIHSS score:** NIHSS scores immediately before randomization, and at 1 day and 7 days post treatment will be collected;
11. **Quality of life using EQ-5D:** EQ-5D quality of life score at 90 days post treatment will be collected.
12. **Inclusion/exclusion criteria evaluation:** Subjects will be evaluated for eligibility using the inclusion and exclusion criteria after signing the informed consent form, to determine whether they can be randomized;
13. **Randomization and thrombectomy treatment:** Subjects will be randomized to the intervention group and the control group at the ratio 1:1 after confirming the eligibility, and receive thrombectomy treatment in the intervention group or control group according to the result of randomization. Information related to thrombectomy treatment will be collected and recorded, which include but not limited to: information of product used for embolectomy, technical success rate, reperfusion outcome in final angiogram and after the first pass, time from femoral artery puncture to successful reperfusion, revascularization quality within 45 minutes, mechanical embolectomy attempts (final), and occurrence of emboli to a new territory, etc.;
14. **Safety evaluation:** All-cause death, intracranial hemorrhage, other SAEs, and procedure-related complications during the study will be collected;
15. **Concomitant medication:** including thrombolytics, anticoagulants, antiplatelets, blood pressure/lipid and blood glucose control medications taken by the subjects during the study, and concomitant medications associated with any SAE.

##### 6.2.2 Study Visits

The study consists of a total of 4 visits, including: Visits at baseline (pre-procedure~ the day of treatment), 1 day (±12 hours) post treatment, 7 days (±2 days) post treatment or at discharge, and 90 days (±14 days) post treatment.

###### 6.2.2.1 Visit 1: Baseline Visit (Pre-operation to the Day of Treatment)

At baseline, subjects will firstly be screened for the study. Each site should keep log of all AIS patients with LVO confirmed by brain imaging arriving <24 hours of symptom onset (or last known well) and who were considered for the study but subsequently excluded. The screening log will record patients' initials and date of admission together with a brief description of the main reason as to why a patient was not randomized. The log will be used by the Research Coordinator, PI and the CCC to monitor recruitment and to identify specific barriers to randomisation of eligible patients. The investigator should introduce the potential subjects or approved surrogates about the purpose of the study and the contents in the informed consent form (ICF). Data collection would begin only when both the subject or his/her surrogate and the investigator have signed and dated on the ICF.

The following information will be collected for subjects at this visit:

1) Date of signature on ICF.
2) **Demographics**: Age, sex, height and weight.
3) **Medical history**: Including medical history of stroke, carotid artery disease, cardiac disorders, hypertension, diabetes mellitus, and hypercholesterolaemia et al, stroke risk factors including history of smoking, alcohol drinking etc, and history of allergy (allergen).
4) **Vital signs**: Pulse, blood pressure, respiration rate, and body temperature.
5) **Laboratory tests:**
   a) Blood routine: Red blood cells (RBC) count, white blood cells (WBC) count, neutrophils count, lymphocytes count, monocytes count, platelets (PLT) count, and hemoglobin (HGB).
   b) Blood biochemistry: Creatinine (Cr), blood urea or blood urea nitrogen (Urea/BUN), and glucose (Glu).
   c) Coagulation function: Prothrombin time (PT), activated partial thromboplastin time (APTT), and international normalized ratio (INR).
6) **Imaging examination:** Data of brain imaging examinations (such as CT/CTA, MRI/MRA or DSA) performed at this visit will be collected.
7) **mRS score:** mRS score of the subjects before this onset will be collected.
8) **Glasgow Coma Scale (GCS) score:** GCS score immediately before randomization will be collected.
9) **NIHSS score:** NIHSS score immediately before randomization will be collected.

10) **Inclusion/exclusion criteria evaluation:** Eligibility of the subjects will be evaluated according to the inclusion and exclusion criteria after obtaining signed informed consent form, to determine whether they can be randomized.

11) **Randomization and thrombectomy treatment:** subjects will be randomized via an Internet-based randomization system in a 1:1 manner to treatment with BGCs(Intervention group) or not(control group), stratified by site, preferred thrombectomy strategy(stent retriever VS aspiration VS stent retriever + aspiration), and onset to randomization time( <6 hours VS ≥6 hours)

    Information related to thrombectomy procedure will be collected and recorded, which include but not limited to: Information of product used for thrombectomy, technical success rate, reperfusion outcome in final angiogram and after the first pass, time from femoral artery puncture to successful reperfusion, revascularization quality within 45 minutes, thrombectomy attempts (final), and occurrence of emboli to a new territory, etc.

12) **Safety evaluation:** All-cause death, intracranial hemorrhage, other SAEs, and procedure or device related complications experienced by subjects during the study will be collected.

13) **Concomitant medication:** Thrombolytics, anticoagulants, antiplatelets, blood pressure/lipid and blood glucose control medications taken by the subjects during the study, and concomitant medications associated with any SAE will be collected.

**6.2.2.2 Visit 2: 1 day (±12 hours) post treatment**

The following information will be collected from the subjects at this visit:

1) Vital signs: Pulse, blood pressure, respiration rate, and body temperature;

2) Laboratory tests:

    a) Blood routine: Red blood cells (RBC) count, white blood cells (WBC) count, neutrophils count, lymphocytes count, monocytes count, platelets (PLT) count, and hemoglobin (HGB);

    b) Blood biochemistry: Creatinine (Cr), blood urea or blood urea nitrogen (Urea/BUN), and glucose (Glu);

c) Coagulation function: Prothrombin time (PT), activated partial thromboplastin time (APTT), and international normalized ratio (INR);

3) Imaging examination: results of imaging examinations such as CT scan will be collected;
4) GCS score;
5) NIHSS score;
6) Safety evaluation: All-cause death, intracranial hemorrhage, other SAEs, and procedure-related complications experienced by subjects during the study will be collected;
7) Concomitant medication: Thrombolytics, anticoagulants, antiplatelets, blood pressure/lipid and blood glucose control medications taken by the subjects during the study, and concomitant medications associated with any SAE will be collected.

###### 6.2.2.3 Visit 3: 7 days (±2 hours) post treatment or at Discharge

The following information will be collected for subjects at this visit:

1) Vital signs: Pulse, blood pressure, respiration rate, and body temperature;
2) GCS score;
3) NIHSS score;
4) Imaging examination: Data of imaging examinations such as CT scan will be collected;
5) Medical cost;
6) Safety evaluation: All-cause death, intracranial hemorrhage, other SAEs, and procedure-related complications experienced by subjects during the study will be collected;
7) Concomitant medication: Thrombolytics, anticoagulants, antiplatelets, blood pressure/lipid and blood glucose control medications taken by the subjects during the study, and concomitant medications associated with any SAE will be collected.

###### 6.2.2.4 Visit 4: 90 days (±14 days) post treatment

1) mRS;
2) Quality of life evaluation using EQ-5D;
3) Medical cost;

4) Safety evaluation: All-cause death, intracranial hemorrhage, other SAEs, and procedure-related complications experienced by subjects during the study will be collected;

5) Concomitant medication: Thrombolytics, anticoagulants, antiplatelets, blood pressure/lipid and blood glucose control medications taken by the subjects during the study, and concomitant medications associated with any SAE will be collected.

### 7 Statistical Analysis

#### 7.1 Sample Size

In a meta-analysis comparing BGC with standard guide catheter, the use of balloon guide catheter improved the proportion of functional outcome by 10.8%[22]. In this study, based on a more conservative estimate, we assume the usage of BGC would improve the function outcome by 8%.

### 8 Study Monitoring

Qualified Clinical Research Associates (CRAs) will be responsible for regular monitoring of this study according to the monitoring plan. The monitoring is to ensure that all parties involved in the clinical study comply with the requirements in the Good Clinical Practice (ICH-GCP/GCP) and other applicable regulations during the implementation of the study, to protect the rights of the subjects and ensure the compliance with the clinical study protocol/procedures, as well as the authenticity, completeness and accuracy of the collected data.

Monitoring procedures include one or more visits before study initiation, regular monitoring visits during the study according to the agreed frequency of monitoring, and monitoring visits related to site closure after the end of the study.

### 9 Data Management

#### 9.1 Data Collection

Electronic data capture (EDC) system will be used in this study, collected data will be entered into the electronic case report form (eCRF) by the investigator or authorized staff, who will receive extensive training prior to data entry. Measures will be taken to ensure the information security.

The investigators are responsible for maintaining all original documents. A complete eCRF for each enrolled subject, regardless of the duration of his/her participation, should be submitted by the investigators or authorized staff. All supporting documents (e.g., laboratory records or records of various clinical research organizations) submitted with the eCRF would be marked with study number, all personal privacy information (including subject names) would be confidential to protect the privacy of the subjects.

##### 9.2 Data Entry and amendment

Investigators and authorized staff would be responsible for data entries, corrections and amendments. Any change to the data will be documented, i.e. reason for change, name of operator, and the time and date of change should be documented. Any correction to the data in the eCRF should signed by the investigators. When there was doubt about the collected data, CRA or data management personnel will raise the query in the EDC system, it is the responsibility of the investigators to answer the query. The EDC system will document the audit trail, including the investigator's name, time and date of the query.

##### 9.3 Lock of Database

After verified by the data management personnel, principal investigator (PI), statistician, and study monitoring personnel, the data administrators will lock the database. Generally, the locked database should not be changed.

##### 9.4 Transfer of Database

The locked database will be transferred to statistician for statistical analysis according to statistical analysis plan, who will develop the statistical analysis report after completing the statistical analysis.

#### 10 Adverse Events/Serious Adverse Events

##### 10.1 Adverse Events (AEs)

###### 10.1.1 Definition of Adverse Events

Adverse events (AE) are any untoward medical occurrence in the clinical investigation, whether or not considered to be related to the device / procedure.

###### 10.1.2 Classification of Adverse Events

**1) Classification by the severity of AE**

The course, severity, treatment and outcome of AEs should be monitored carefully and recorded in the AE reporting form. The severity of an AE can be classified as mild, moderate, or severe according to the following criteria.

**Table 10-1 Adverse Events by Severity**

| Severity | Description |
|---|---|
| Mild | Transient or mild discomfort, with no limitation of activity, and no medical intervention/treatment required. |
| Moderate | Obvious limitation of activity, often requiring assistance, medical intervention/treatment, and hospitalization may be necessary. |
| Severe | Extreme limitation of activity, requiring significant assistance and significant medical intervention/treatment, and requiring hospitalization or probably requiring hospice care. |

**2) Classification by relationship of AE to the study device/procedure:**

For all collected AEs, the relationship of the AE to the study device or procedure will be determined by the investigator based on temporal relationship and his/her clinical judgment. The following categories will be used for grading relevance.

**Criteria for a causal relationship of AE and the Study Device or Procedure:**

1) AE follows a fair temporal sequence from the study device or procedure;
2) The event is a known effect of the study device or the procedure, or a possible effect can be explained by the mechanism of the study device or the procedure;
3) The event relieves or ceases after stopping use of the device or procedure;
4) The event reappears after reuse of the device or procedure;
5) The event could not be explained by other contributing factors.

Any event meeting five items of the criteria above will be considered as "definitely related"; while any event meeting two of the criteria above will be considered as "possibly related".

**Criteria for a relationship unrelated to the Study Device or Procedure:**

1) AE follows a fair temporal sequence from the study device or procedure ;
2) This type of adverse events is impossible to be caused by the study device or surgical procedure;

3) The adverse event can be explained by the concomitant use of other device/drug, subject's disease progression, or other treatment effects.

Any event meeting three items of the criteria above will be considered as "definitely unrelated"; while any event meeting one of the criteria above will be considered as "unlikely related".

Among which, events whose causal relationship is judged as definitely related or possibly related will be considered as related to the study device or procedure.

###### 10.1.3 Recording and Follow up of Adverse Events

The investigator or authorized staff will document serious adverse events and procedure-related complications occurring at any time during the study after the subject has signed the informed consent form. Any serious adverse event or procedure-related complication should be treated and followed up until the event has been reasonably resolved (e.g. symptoms recovered, resolved, or reach to a stable state, or can be well-explained). All treatments and results for SAEs and procedure-related complications must be documented appropriately on the CRF.

##### 10.2 Serious Adverse Event (SAE)

###### 10.2.1 Definition of Serious Adverse Events

Any serious adverse event (SAE) that occurs during the study will be recorded by the investigator from the time that the study treatment is started for the subject. SAE reporting and notification requirements are based on the International Conference on Harmonization-Good Clinical Practice (ICH-GCP) guidelines. According to the WHO International Centre for Drug Monitoring (1994), a SAE is defined as any of the following untoward medical events which:

1) Results in death;
2) Is life threatening in the view of the principal investigator (i.e. its occurrence places the patient or subject at immediate risk of death. It does not include an adverse event, had it occurred in a more severe form, might have caused death);
3) Requires inpatient hospitalization or prolongation of existing hospitalization;
4) Results in a persistent or significant disability or incapacity;
5) Result in congenital anomaly or birth defects;
6) Important medical events that may not result in death, be life-threatening, or require hospitalizations but may jeopardize the subject and may require medical or surgical

intervention to prevent one of the outcomes listed above according to the principal investigator.

Notes: Any hospital admission planned due to existed illness or disease before enrolment, or as required by the study when the subjects did not experience health deterioration, are not considered as a SAE.

##### 10.2.2 Reporting of Serious Adverse Events

In case of a SAE, the investigator should immediately take appropriate treatment for the subject; meanwhile the investigator should report the SAE, within 24 hours after awaring of it, to the ethics committee and the sponsor, and follow up the SAE as required in the protocol and submit follow-up and summary reports as required.

### 11 Ethical Considerations

#### 11.1 Ethical Requirements in Relevant Regulations

This clinical study must be conducted in compliance with Declaration of Helsinki, and other applicable regulations. The study can be initiated only after the files such as the study protocol and ICF have been approved by the ethics committee of the site.

For every subject, before being enrolled in this study, the investigator is responsible for giving him/her or his/her surrogate a complete and comprehensive introduction to the objectives, procedures and possible risks of this study, and they should be made aware that they can withdraw from the study at any time. Then, the ICF will be signed by the subject or his/her surrogate and the investigator, and the signed ICF should be retained as a clinical study document for inspection.

Before initiation of this clinical study, the investigator should submit this clinical study protocol, ICF, and other relevant documents to the ethics committee. This study can only be initiated after being approved by the Ethics Committee. Any amendment to the protocol must be approved by the Ethics Committee before implementation. Any SAE occurring during the clinical study should be reported to the Ethics Committee in writing within 24 hours after the investigator's awareness of the event.

Before the start of the study in each clinical trial institution, the Ethics Committee of the principal sites are responsible for reviewing the ethical appropriateness, rationality and scientificity of the study protocol. The ethics committees of other sites should review feasibility of the study in their institutions by means of conference or document review, when accepting the review comments of the ethics committee of the principal site, including the qualification and experience of the investigators, study equipments and conditions, etc.

The ethics committees shall supervise the clinical study in their corresponding clinical trial institutions, and may require to suspend or terminate the study at any time via written notification if it finds that the subjects' rights and interests cannot be guaranteed.

The clinical study, once suspended, shall not be resumed without the approval of the Ethics Committee.

Should any SAE occur in this study, the investigator should immediately take appropriate treatment measures for the subject; meanwhile a written report shall be submitted to the Ethics Committee of the site.

##### 11.2 Informed Consent

Before a subject agrees to participate, the investigator should fully explain the details of this study, including the purpose, contents, and procedures as well as the possible risks and benefits of this study, to the subject or the surrogate and/or the impartial witness (according to local regulations). After a full and detailed explanation, the ICF will be signed and dated by the subject or his/her surrogate and/or the impartial witness (according to local regulations) as well as the investigator.

The ICF will be signed in duplicates by the investigator and the subject or his/her guardian and/or a third-party witness, with each keeping one copy.

##### 11.3 Approval of the Study Protocol

The ethics committee is responsible for the review and approval of this clinical study protocol, and the study could not be initiated until final approval is obtained from the ethics committee.

#### 12 Quality Control of the Study

##### 12.1 Investigators Training

Prior to the start of the study, the investigators will be trained on the study protocol by a CRA designated by the sponsor.

##### 12.2 Measures to Improve Subjects Compliance

The investigator should conduct informed consent carefully to ensure that the subjects fully understand the study requirements and cooperate during the trial.

##### 12.3 Clinical Monitoring

The CRAs designated by the sponsor will perform on-site monitoring visits regularly to ensure that the study protocol is strictly followed and to check the consistency of the contents in the eCRF with the source data.

##### 12.4 Data Management

The eCRF should be completed as soon as possible during or after the visit and updated timely to ensure that it reflects the latest status of the subjects in the study. The investigator should review the data to ensure the accuracy and correctness of all data entered into the eCRF. If some assessments are not performed during the study, or some information is unavailable, not applicable, or unknown, the investigator should also record it in the eCRF. The investigator shall electronically sign on the completed and verified CRF/the verified data.

##### 12.5 Maintenance of Source Documents

Source documents refer to all documents of the subjects used by the investigator or the clinical study institution, which can prove the existence of the subjects, the inclusion and exclusion criteria, and records for their participation in this study, including laboratory records, and subject folder, etc.

The clinical study institution, investigator and sponsors shall establish a retention system of essential documents. The study essential documents can be divided into three parts based on the phase of the study, i.e. files in the preparing phase, ongoing phase, and termination or completion phase. The clinical study institution should properly retain the study records and essential documents as agreed with the sponsor.

Source documents for this study include signed informed consent forms, laboratory test reports, operation records of each subject, and other relevant records.

#### 13 Study Administration

##### 13.1 Organizational Structure

The study consists of a steering committee (SC) of experts in the therapeutic field who are responsible for the oversight of the study. CCRC Medtech (Shanghai) Co., Ltd. will serve as the central coordinating center (CCC), to be responsible for the organization and operation of the study. The specific work of the committee is described below.

##### 13.1.1 Steering Committee

**Responsibilities:** Responsible for the overall study design, protocol implementation, data collection, analysis plan and subsequent publication. The Committee can appoint new members and co-elect other members to enhance the integrity of the research and analysis work.

**Members:** Core members of the steering committee are listed below and other members will be specified in the charter of the committee.

- Professor Jianmin Liu (Co-Principal Investigator), The First Affiliated Hospital of Naval Medical University（Changhai hospital）, China;
- Professor Mayank Goyal (Co-Principal Investigator), University of Calgary Cumming School of Medicine.
- Professor Pengfei Yang, The First Affiliated Hospital of Naval Medical University（Changhai hospital）, China;

##### 13.1.2 Central Coordinating Center (CCC)

CCRC Medtech (Shanghai) Co., Ltd. will serve as the CCC.

Responsibilities of CCC: Daily administration of the study, data and project management, committee coordination, assistance in applications to ethics committees, training of participating sites on the study protocol and procedures, site initiation visits, monitoring of data quality and compliance with applicable guidelines and regulations.

##### 13.1.3 Imaging Adjudication Committee (Core Laboratory)

Responsibilities: De-identification of images, and independent assessment of all imaging data of CT/CTA, MRI/MRA, and DSA.

##### 13.1.4 Clinical Events Committee (CEC)

Responsibilities: Review the SAEs and procedure related complications, to ensure the criteria for evaluation are consistent.

###### 13.1.5 Data and Safety Monitoring Board (DSMB)

Responsibility: Review the safety, ethicality and study results throughout the study.

###### 13.1.6 Outcome Assessment Committee (OAC)

Responsibilities: independent assessment of the primary outcome (mRS score) in blinded manner.

##### 13.2 List of Study Sites

The study, leading by The First Affiliated Hospital of Naval Medical University（Changhai hospital）, is planned to be conducted at 40-60 sites in China. A list of all participating sites will be provided separately.

##### 13.3 Confidentiality Principle

This protocol is a confidential document provided to the study team members including the medical experts and investigators in this study, as well as medical institutions, ethics committees, CROs and other concerned institutions undertaking this trial. Except for explaining relevant information to the subjects, no part of this protocol can be made public or disclosed to any third party without prior written consent of the sponsor. In addition, the results of this study, regardless of in part or as a whole, can only be presented to any society or published in any journal after receiving written consent of the sponsor.

The clinical study agreement, the contents of this clinical study and all supplementary materials are confidential and exclusively owned by the sponsor. The investigator should keep confidential of the information including patent application, manufacturing process and unpublished data provided by the sponsor, etc., and shall not disclose any information to the third party unless agreed by the sponsor. This confidentiality obligation remains effective after the termination or end of this study.

The personal data of the subjects collected by the investigator and his/her staff for use in this study (“study data"), include date of birth, sex, identity card, home address, photographs, and personal data regarding physical or mental health status of the subjects.

All medical records and study materials that can identify the subjects should be kept confidential to the extent permitted by law. However, the investigator, the sponsor and its representatives, the CRAs (the persons responsible for monitoring how the study is conducted and ensuring that the information is collected correctly), and in certain specific circumstances, regulatory authorities and ethics committees may inspect and copy confidential information that identifies the subjects. All personal information in the study will be handled in accordance with the national and local laws on data protection.

Subjects have the right to request the investigator and the sponsor to protect their information and to correct any inaccuracies in these data. If a subject withdraws consent, the investigator will no longer use or disclose the subject's data to others, but the sponsor may still use the study data obtained before withdrawal.

The results of this study may be published in medical journals and presented at medical meetings. Subjects will not be identified in any of these publications.

##### 13.4 Agreement on the Publication of Study Results

The main results of this study will be published in the name of the PROTECT-MT Collaborative Research Group. Publication of the results will be managed and controlled primarily by a writing committee designated by the SC.

The investigator can publish or present the study results. However, as this is a multi-center study, the investigator must agree that: interim results of the study may not be published or presented without prior written permission of the SC. In addition, the investigator must agree that: notify SC at least 30 days prior to submission of the study results for publication or presentation, and provide a summary of the study results report or a copy of the manuscript (including but not limited to text, PPT slides, and any other translated text or media materials). SC shall review and provide comments on the publications, abstracts, slides and manuscripts; review and comment on the accuracy of the information; protect the rights of any individual; ensure fairness and compliance with the applicable regulations.

In the event of disagreement between the parties regarding the appropriateness and/or confidentiality of the data analysis and presentation, the investigator shall agree to meet with SC members at the study site or as otherwise agreed to discuss and resolve any disagreement prior to submission for publication.

#### 14 Fund

This study is funded by Shanghai Shenkang Hospital Development Center , Biopharma Industry Promotion Center Shanghai and Zhuhai TonBridge Medical Technology Co., Ltd.

### 15 Appendix

#### Appendix 1: NIHSS Scale
